# Genomic epidemiology and antimicrobial resistance transmission of *Salmonella* Typhi and Paratyphi A at three urban sites in Africa and Asia

**DOI:** 10.1101/2023.03.11.23286741

**Authors:** Zoe A. Dyson, Philip M. Ashton, Farhana Khanam, Angeziwa Chunga, Mila Shakya, James Meiring, Susan Tonks, Abhilasha Karkey, Chisomo Msefula, John D. Clemens, Sarah J. Dunstan, Stephen Baker, Gordon Dougan, Virginia E. Pitzer, Buddha Basnyat, Firdausi Qadri, Robert S. Heyderman, Melita A. Gordon, Andrew J. Pollard, Kathryn E. Holt, STRATAA Study Group

**Author notes:** Corresponding authors: Zoe A. Dyson, Kathryn E. Holt. These authors contributed equally to this work (Philip M. Ashton, Farhana Khanam, Angeziwa Chunga, Mila Shakya, James Meiring). These authors contributed equally to this work (Buddha Basnyat, Firdausi Qadri, Robert S. Heyderman, Melita A. Gordon, Andrew J. Pollard).

## Abstract

**Background:** Enteric fever is a serious public health concern. The causative agents, *Salmonella enterica* serovars Typhi and Paratyphi A, are frequently antimicrobial resistant (AMR), leading to limited treatment options and poorer clinical outcomes. We investigated the genomic epidemiology, resistance mechanisms and transmission dynamics of these pathogens at three urban sites in Africa and Asia.

**Methods:** Bacteria isolated from febrile children and adults at study sites in Dhaka, Kathmandu, and Blantyre were sequenced and AMR determinants identified. Phylogenomic analyses incorporating globally-representative genome data, and ancestral state reconstruction, were used to differentiate locally-circulating from imported pathogen variants.

**Findings:** *S*. Paratyphi A was present in Dhaka and Kathmandu but not Blantyre. *S*. Typhi genotype 4.3.1 (H58) was common in all sites, but with different dominant variants (4.3.1.1.EA1 in Blantyre; 4.3.1.1 in Dhaka; 4.3.1.2 in Kathmandu). Resistance to first-line antimicrobials was common in Blantyre (98%) and Dhaka (32%) but not Kathmandu (1.4%). Quinolone-resistance mutations were common in Dhaka (99.8%) and Kathmandu (89%) but not Blantyre (2.1%). *AcrB* azithromycin-resistance mutations were rare (Dhaka only; n=5, 1.1%). Phylogenetic analyses showed that (a) most cases derived from pre-existing, locally-established pathogen variants; (b) nearly all (98%) drug-resistant infections resulted from local circulation of AMR variants, not imported variants or recent *de novo* emergence; (c) pathogen variants circulated across age groups. Most cases (67%) clustered with others that were indistinguishable by point mutations; individual clusters included multiple age groups and persisted for up to 2.3 years, and AMR determinants were invariant within clusters.

**Interpretation:** Enteric fever was associated with locally-established pathogen variants that circulate across age groups. AMR infections resulted from local transmission of resistant strains. These results form a baseline against which to monitor the impacts of control measures.

**Funding:** Wellcome Trust, Bill & Melinda Gates Foundation, European Union’s Horizon 2020, NIHR.

**Research in context:** *Evidence before this study:* Current knowledge of the enteric fever pathogen populations in Dhaka, Kathmandu, and Blantyre comes from retrospective analysis of isolates captured from routine diagnostics or treatment trials. Due to these study designs, most focus on either adult or paediatric cohorts, which complicates assessment of pathogen variant transmission across age groups. Many studies report prevalence of antimicrobial resistance (AMR) and associated mechanisms amongst enteric fever cases. Genomic studies at these sites and elsewhere have identified the spread of AMR clones, and a recent genomic study quantified the inter- and intra-continental spread of resistant *S*. Typhi between countries. However, PubMed search of “(typhoid OR (enteric fever)) AND (genom*)” identified no studies quantifying the relative proportion of resistant infections that is attributable to local transmission of resistant variants vs imported strains or *de novo* emergence of AMR.

*Added value of this study:* We estimate the vast majority (98%) of drug-resistant enteric fever cases identified in our study resulted from local circulation of resistant variants. Further, we show genetically indistinguishable pathogen variants (either resistant or susceptible) persisting for up to 2.3 years and causing infections across all age groups (under 5 years; 5-15 years; ≥15 years).

*Implications of all the available evidence:* While inter-country transfer of resistant enteric fever pathogens does occur and is concerning, the burden of drug-resistant enteric fever at the study sites is currently caused mainly by transmission of locally-established variants, and transmits across age groups. These data confirm assumptions made in models of vaccine impact regarding heterogeneity of pathogen variants and AMR across age groups, and support that childhood immunisation programmes can be expected to reduce the overall burden of resistant infections in endemic settings.

## Introduction

The global burden of enteric fever is estimated at 14.3 million cases annually^1^ and is concentrated in South Asia and Sub-Saharan Africa. The causative agents are *Salmonella enterica* serovars Typhi and Paratyphi A (*S*. Typhi and *S*. Paratyphi A), though the latter is rarely identified in Africa. Mortality and complications occur at significantly higher rates in the absence of effective antimicrobial therapy. Antimicrobial resistant (AMR) pathogen variants are common, but the specific drugs and causative mechanisms vary widely. Most regions have seen a decline in resistance to the older first-line drugs (chloramphenicol, co- trimoxazole and ampicillin; the combination of which is defined as multi-drug resistant, MDR) and an increase in fluoroquinolone non-susceptibility (ciprofloxacin MIC ≥0.06 mg/L)^2, 3^. The emergence of resistance to commonly-used oral agents cefixime and azithromycin has been reported sporadically, mainly in South Asia^4–7^, and extensively-drug resistant (XDR, defined as MDR plus resistant to ciprofloxacin and third-generation cephalosporins) *S*. Typhi is now common in Pakistan^4^. Both regional transfer and local clonal expansion of AMR variants have been documented^7–10^, but the extent to which AMR disease burden is attributable to transmission of endemic AMR variants, versus importation of exogenous AMR variants or *de novo* emergence of AMR in the local population, has not been specifically quantified in settings with endemic disease.

Vaccines against *S*. Typhi infection have been used in travellers for decades, but mass immunisation programmes have not been applied in most endemic areas. Recently-licensed Gavi-supported typhoid conjugate vaccines (TCVs) offer new opportunities to reduce the burden of *S*. Typhi infection^11^. Trials have shown these vaccines to be safe and immunogenic in children (from 9 months of age), with >80% efficacy against incident *S.* Typhi infection^12–15^. In Pakistan and Zimbabwe, TCV immunisation campaigns have been deployed to help control XDR and ciprofloxacin-resistant *S*. Typhi outbreaks, respectively^16, 17^. These successful campaigns were followed by introduction of national typhoid immunisation programmes^18, 19^, which are now being considered by several other countries.

It is important to monitor the impact of new vaccine programmes on the underlying pathogen populations. TCVs do not cross-protect against *S*. Paratyphi A infection, and *S*. Paratyphi A may expand to fill the niche. TCVs are effective against some AMR *S*. Typhi^18, 19^, but it is unknown whether they will be equally effective against all *S*. Typhi variants, or promote emergence of vaccine-escape mutants or AMR variants. Baseline data on pathogen populations is therefore essential; whole-genome sequencing (WGS) is now the standard as it provides high-resolution data simultaneously on lineage diversity, resistance mechanisms and transmission dynamics. We recently assessed the enteric fever burden in three urban sites (Blantyre, Malawi; Kathmandu, Nepal; Dhaka, Bangladesh) as part of the Strategic Typhoid Alliance across Africa and Asia (STRATAA)^20^. Current knowledge of the enteric fever pathogen populations at STRATAA sites comes from retrospective analysis of isolates captured from routine diagnostics^3^ or treatment trials^10^ conducted using different protocols, and usually separated into adult or paediatric focused studies^3, 10^. The available data indicate that *S*. Typhi H58 (subclade 4.3.1 and derived genotypes) has been dominant across South Asia as well as Eastern and Southern Africa, including the STRATAA sites, for many years^2, 3, 8–10^. In Malawi, *S*. Typhi epidemics have been documented since the late 1990s and are now associated almost entirely with MDR genotype 4.3.1, which has been clonally expanding since its arrival in 2009; the disease is now considered endemic^8, 9^. In contrast, in Bangladesh and Nepal, *S*. Typhi and *S*. Paratyphi A have been hyperendemic for decades and probably centuries; this is reflected in a diversity of co-circulating pathogen genotypes, although the populations have been dominated by 4.3.1 for the last two decades^2, 3, 21^.

Here we use WGS to investigate the pathogen populations underlying enteric fever at STRATAA sites, collected prospectively in defined catchment areas using the same protocol, providing valuable baseline data ahead of vaccine trials^12–15^ and planned national immunisation programmes^19^. We characterise the pathogen populations and AMR determinants, investigate transmission patterns across age groups, and quantify the proportion of AMR cases that are attributable to local transmission of endemic AMR variants.

## Results

### Pathogen diversity and population structure

We sequenced n=731 unique typhoidal *Salmonella* blood-culture isolates from febrile individuals recruited in the STRATAA catchment areas during passive surveillance for enteric fever^20, 22^ (see **Methods, Tables 1 and S1**). The distribution of serotypes and genotypes is shown in **Fig. 1**. Genomic analysis confirmed^20^ that serovar Paratyphi A was present in both the South Asian sites (n=95, 21.0% of sequenced cases in Dhaka; n=14, 10.1% of sequenced cases in Kathmandu) but not in Blantyre. Across all three sites, *S*. Typhi 4.3.1 (H58) genotypes were most frequently observed, representing between 42.3-97.9% of sequenced enteric fever cases (**Table 1**). However, different H58 subtypes were present at each of the three sites, which also differed markedly in terms of the overall diversity of pathogen variants causing enteric fever (**Fig. 1, Table 1**). Enteric fever cases in Blantyre were caused almost exclusively by *S.* Typhi 4.3.1.1.EA1 (97.9%), and thus displayed very low genotype diversity (Simpson’s index = 0.05). Enteric fever cases in Kathmandu were more diverse (Simpson’s index = 0.67), with one-third caused by a high abundance non-H58 *S*. Typhi genotype (3.3.2) and one-half by *S*. Typhi 4.3.1.2 (**Fig. 1, Table 1**). Dhaka was the most diverse setting (Simpson’s index = 0.80), with two common lineages of H58 *S*. Typhi (genotypes 4.3.1.1, 39.2% and 4.3.1.3.Bdq, 2.7%), four non-H58 *S*. Typhi genotypes with ≥25 sequenced cases each (2.0.1, 2.1.7, 2.3.3, 3.2.2), and two common *S*. Paratyphi A genotypes each associated with ≥20 sequenced cases (2.3.3 and 2.4.4) (**Fig. 1, Table 1**).

**Figure 1.**
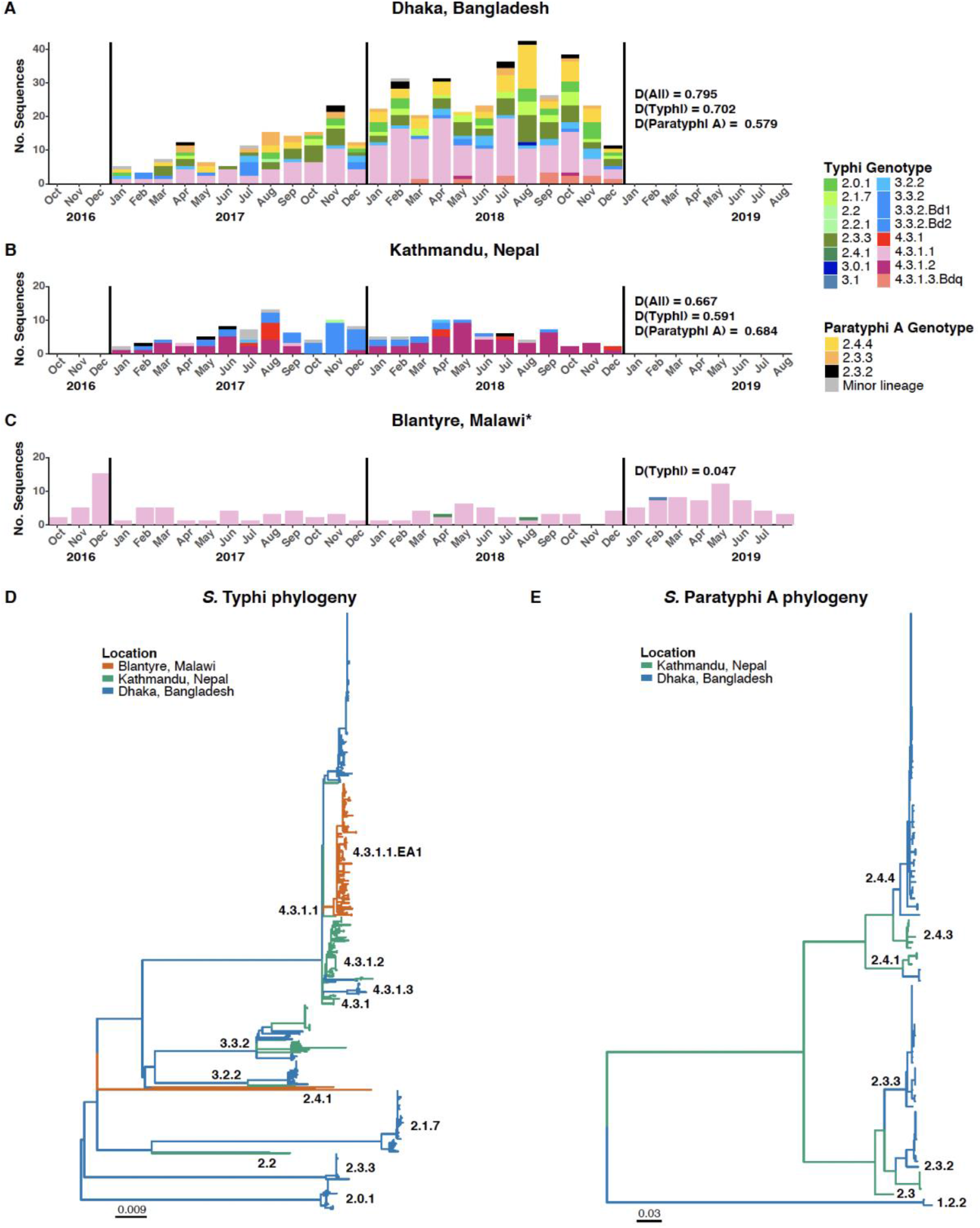
Genetic diversity of enteric fever pathogen populations. **(A-C)** Epidemic curves for sequenced enteric fever cases in the three STRATAA catchment areas, stratified by pathogen genotype. Bars indicate monthly case counts, coloured to indicate serovar and genotype as per inset legend. *4.3.1.1 in Blantyre is sublineage 4.3.1.1.EA1. Inset labels ‘D( )’ indicate Simpson’s diversity index calculated from pathogen genotype counts for *S.* Typhi, *S.* Paratyphi A or both (‘D(All)’). **(D-E)** Maximum-likelihood phylogenetic trees for STRATAA isolates of serovars Typhi (D) and Paratyphi A (E), inferred from genome-wide single nucleotide variant alignments. Each tree was outgroup-rooted using genomes of the other serovar. Branches are coloured by country as per inset legend, and labelled by genotype. Data in all panels (A-E) represent the main STRATAA dataset (n=622 *S*. Typhi and n=109 *S*. Paratyphi A; see Table 1).

**Table 1.** Summary of enteric fever cases and sequenced pathogens. For each serovar (Typhi, Paratyphi A), the table shows total sequenced cases; number and frequency of common genotypes (accounting for ≥10% total sequenced cases in at least one site); and Simpson’s diversity index (calculated from genotype counts). *Blantyre had an extended surveillance period; data from the full period (34 months) is included in the main STRATAA dataset used for phylogenetic and statistical analyses (n=622 *S*. Typhi and n=109 *S*. Paratyphi A), however the first 24-month period was used to calculate Simpson’s diversity so as to be comparable across sites.

|  | **Dhaka** | **Kathmandu** | **Blantyre** |  |
| --- | --- | --- | --- | --- |
| Surveillance period | 24 months<br>(Jan 2017–Dec 2018) | 24 months<br>(Jan 2017–Dec 2018) | 24 months<br>(Oct 2016–Sep 2018) | *34 months<br>(Oct 2016–Aug 2019) |
| Unique cases recruited, N | 454 | 164 | 115 | 158 |
| Unique cases sequenced, N (%) | 452 (99.6%) | 138 (84.1%) | 83 (72.2%) | 141 (89.2%) |
| **Characteristics of sequenced cases** |  |  |  |  |
| Male, N (%) | 240 (53.1%) | 82 (59.4%) | 33 (39.8%) | 64 (45.4%) |
| <5 years, N (%) | 105 (23.2%) | 10 (7.3%) | 17 (20.5%) | 27 (19.1%) |
| ≥5 and <15 years, N (%) | 208 (46.0%) | 60 (43.5%) | 48 (57.8%) | 72 (51.1%) |
| ≥15 years, N (%) | 139 (30.8%) | 68 (49.3%) | 18 (21.7%) | 42 (29.8%) |
| **S. Typhi** |  |  |  |  |
| Total sequences, N (%) | 357 (79.0%) | 124 (89.9%) | 83 (100%) | 141 (100%) |
| 4.3.1 (H58), N (% of serovar) | 191 (53.5%) | 80 (64.5%) | 81 (97.6%) | 138 (97.9%) |
| 4.3.1.1, N (% of serovar) | 177 (49.6%) | 3 (2.4%) | 81 (97.6%) | 138 (97.9%) |
| 4.3.1.2, N (% of serovar) | 2 (0.56%) | 67 (54.0%) | 0 | 0 |
| 2.3.3, N (% of serovar) | 64 (17.9%) | 0 | 0 | 0 |
| 3.3.2, N (% of serovar) | 17 (4.8%) | 41 (33.1%) | 0 | 0 |
| Simpson's diversity | 0.70 | 0.59 | 0.05 | - |
| **S. Paratyphi A** |  |  |  |  |
| Total sequences, N (%) | 95 (21.0%) | 14 (10.1%) | 0 | 0 |
| 2.3.2, N (% of serovar) | 11 (11.6%) | 4 (28.6%) |  |  |
| 2.3.3, N (% of serovar) | 23 (24.2%) | 0 |  |  |
| 2.4.1, N (% of serovar) | 3 (3.1%) | 3 (21.4%) |  |  |
| 2.4.3, N (% of serovar) | 0 | 6 (42.9%) | 0 | 0 |
| 2.4.4, N (% of serovar) | 56 (58.9%) | 0 | 0 | 0 |
| Simpson's diversity | 0.58 | 0.68 | - | - |

We considered three age groups that reflect broadly different contact networks: pre-school age (under 5 years and interacting mainly with other household members), school-age children (5-15 years, interacting with household members and other school-age children), and working age (≥15 years old, interacting with household members and the wider community). In all settings, all three age groups were infected with a diverse range of *S*. Typhi and *S*. Paratyphi A genotypes (**Fig. 2a**). Similarly, at the sub-genotype level, the age groups were intermingled in the maximum-phylogenetic trees (**Figs. S4-6**), with no evidence of clustering by patient age group (*K*=7.16x10^-6-2^.24x10^-5^, where *K*<1 indicates no signal). Within each site, the local prevalence of *S*. Paratyphi A, and of H58 amongst *S*. Typhi, was similar across age groups and sexes (**Fig. 2a**). In Dhaka, where the diversity of serovars and genotypes was greatest, there was a significant association between increasing age groups and prevalence of *S*. Paratyphi A (OR 1.93, [95% CI, 1.39-2.71], p=1x10^-4^ using logistic regression; **Fig. 2b**), and of non-H58 *S*. Typhi genotypes (OR 1.5 [95% CI, 1.08-1.95], p=0.01; **Fig. S1a**) (see **Table S2**). Across all sites, n=93 enteric fever cases were considered severe (see **Methods**); these were all caused by *S.* Typhi and were less common in Kathmandu (n=13, 10.5% vs n=62, 17.4% in Dhaka and n=18, 12.8% in Blantyre; see **Fig. 2c, Table S3**). Severe disease was not significantly associated with patient age-group or sex, *S*. Typhi genotype (H58 vs other) or MDR using logistic regression models (**Fig. 2b**, **Table S3**).

**Figure 2.**
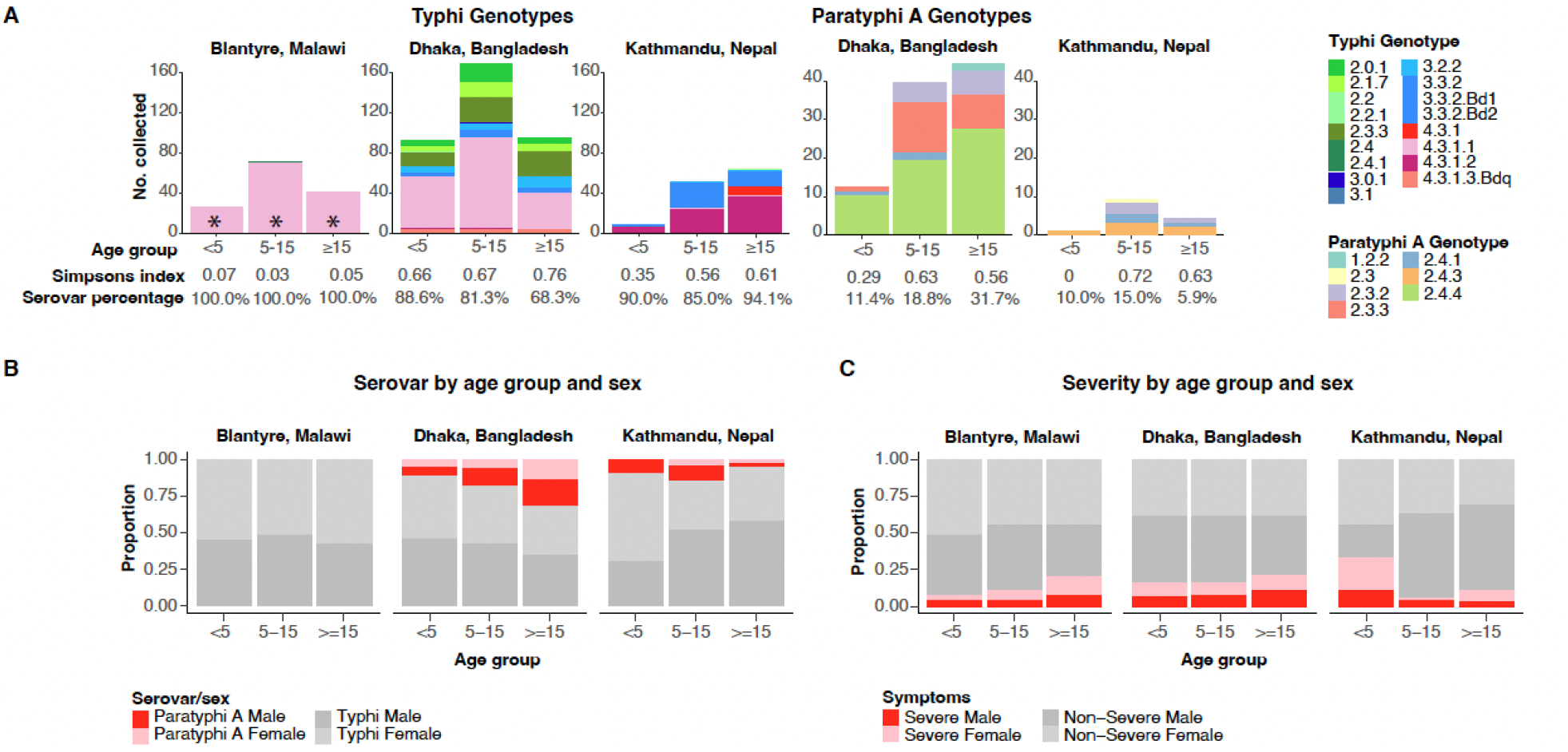
Distribution of pathogen variants and disease severity by age group. (A) Barplots show frequency distributions of pathogen genotypes among age groups, stratified by serovar and location, for the main STRATAA dataset (n=622 *S*. Typhi and n=109 *S*. Paratyphi A; see Table 1). **\***4.3.1.1 in Blantyre is sublineage 4.3.1.1.EA1. Simpson’s diversity index is shown under each bar (calculated from genotype counts, excluding isolates from the extended surveillance period in Blantyre to ensure comparability between locations). Serovar frequencies, amongst all sequenced isolates from the given location and age-group, are also shown under each bar. **(B)** Breakdown of patient sex and pathogen serovar, within each age-group, at each site, for the main STRATAA dataset. **(C)** Breakdown of patient sex and disease severity (severe defined as symptom duration >10 days and/or requiring hospitalisation), within each age-group, at each site, for the main STRATAA dataset.

At each STRATAA site, all local pathogen variants co-circulated throughout the period of surveillance (**Fig. 1**). The dominant genotypes detected in each setting matched those identified in earlier studies^2, 3, 9^, and contextualisation with global genomes supports that most cases derive from locally-established pathogen variants that are now endemic in their respective settings (**Fig. S2**). The exception to this was a cluster of genotype 3.3.2 in Kathmandu, which appears to have been imported from elsewhere in South Asia, with ancestral sequences isolated from Bangladesh (**Fig. S3a**). This 3.3.2 cluster was first isolated in the Kathmandu STRATAA catchment in February 2017 and increased in numbers, peaking in November and December 2017 (85-90% of monthly *S*. Typhi cases, n=9-6 per month), before declining (**Fig. S3b**).

### Antimicrobial resistance

Genetic mechanisms of resistance are summarised in **Table 2**. Acquired resistance genes for first-line drugs, associated with MDR, were detected in *S.* Typhi 4.3.1.1 from Blantyre and Dhaka. Plasmid-borne quinolone-resistance gene *qnrS* was detected in *S.* Typhi 4.3.1.3.Bdq in Dhaka, and mutations in the quinolone resistance determining region (QRDR, associated with non-susceptibility to fluoroquinolones) were common across all *S.* Typhi and *S*. Paratyphi A genotypes in Dhaka and Kathmandu but present in just n=3 4.3.1.1.EA1 in Blantyre. Azithromycin resistance-associated mutation *acrB*-R717Q was detected in two *S.* Typhi and three *S*. Paratyphi A from Dhaka.

**Table 2.** Distribution of AMR determinants. Data shown represents the main STRATAA dataset (n=622 *S*. Typhi and n=109 *S*. Paratyphi A; see Table 1). Genotypes with ≥10 samples, or representing a major 4.3.1 (H58) sublineage, are shown; the remaining genotypes per site are grouped as ‘Rare genotypes’. ‘QRDR’, quinolone resistance determining region; ‘1 QRDR’ indicates one such mutation (associated with decreased ciprofloxacin susceptibility); ‘3 QRDR’ indicates three such mutations (associated with ciprofloxacin resistance). *All acquired genes for first-line drugs were associated with chromosomally integrated transposons ^§^4.3.1.1.Bdq genomes carried *blaTEM-1, sul2, tet(A)* together with *qnrS* in an IncFIB_K_ plasmid.

| Pathogen variant | Frequency<br>N (% of serovar) | Acquired genes*<br>for first-line drugs<br>N (% of genotype) | QRDR & *qnrS*<sup>§</sup><br>(fluoroquinolones)<br>N (% of genotype) | AcrB-R717Q<br>(azithromycin)<br>N (% of genotype) | No AMR<br>N (% of genotype) |
| --- | --- | --- | --- | --- | --- |
| **Blantyre** |  |  |  |  |  |
| *S. Typhi* | 141 | 138 (98%) | 1 QRDR: 3 (2%) | 0 | 3 (2%) |
| 4.3.1.1.EA1 | 138 (98%) | 138 (100%) | 1 QRDR: 3 (2.2%) | 0 | 0 |
| Rare genotypes | 3 (2%) | 0 | 0 | 0 | 3 (100%) |
| **Dhaka** |  |  |  |  |  |
| *S. Typhi* | 357 | 170 (47.6%) | 356 (99.7%) | 2 (0.6%) | 1 (0.3%) |
| 2.0.1 | 31 (8.7%) | 0 | 1 QRDR: 31 (100%) | 0 | 0 |
| 2.1.7 | 27 (7.6%) | 0 | 1 QRDR: 27 (100%) | 0 | 0 |
| 2.3.3 | 64 (17.9%) | 0 | 1 QRDR: 64 (100%) | 0 | 0 |
| 3.2.2 | 25 (7%) | 0 | 1 QRDR: 25 (100%) | 1 (4.0%) | 0 |
| 4.3.1.1 | 177 (49.6%) | 170 (96%) | 1 QRDR: 177 (100%) | 1 (0.6%) | 0 |
| 4.3.1.2 | 2 (0.6%) | 0 | 1 QRDR: 1 (50%)<br>3 QRDR: 1 (50%) | 0 | 0 |
| 4.3.1.3.Bdq | 12 (3.4%) | 12 (100%)<sup>§</sup> | 1 QRDR plus *qnrS*<sup>§</sup>:<br>12 (100%) | 0 | 0 |
| Rare genotypes | 19 (5.3%) | 0 | 1 QRDR: 18 (94.7%) | 0 | 1 (5.3%) |
| *S. Paratyphi A* | 95 | 0 | 95 (100%) | 3 (3.2%) | 0 |
| 2.3.2 | 11 (11.6%) | 0 | 1 QRDR: 11 (100%) | 0 | 0 |
| 2.3.3 | 23 (24.2%) | 0 | 1 QRDR: 23 (100%) | 0 | 0 |
| 2.4.4 | 56 (58.9%) | 0 | 1 QRDR: 56 (100%) | 3 (5.4%) | 0 |
| Rare lineages | 5 (5.3%) | 0 | 1 QRDR: 5 (100%) | 0 | 0 |
| **Kathmandu** |  |  |  |  |  |
| *S. Typhi* | 124 | 2 (1.6%) | 109 (87.9%) | 0 | 14 (11.3%) |
| 3.3.2 | 41 (33.1%) | 0 | 1 QRDR: 31 (75.6%) | 0 | 10 (24.4%) |
| 4.3.1 | 10 (8.1%) | 0 | 1 QRDR: 10 (100%) | 0 | 0 |
| 4.3.1.1 | 3 (2.4%) | 2 (66.7%) | 1 QRDR: 2 (66.7%) | 0 | 0 |
| 4.3.1.2 | 67 (54.0%) | 0 | 1 QRDR: 63 (94%)<br>3 QRDR: 3 (4.5%) | 0 | 1 (1.5%) |
| Rare genotypes | 3 (2.4%) | 0 | 0 | 0 | 3 (100%) |
| *S. Paratyphi A* | 14 | 0 | 1 QRDR: 14 (100%) | 0 | 0 |
| Rare lineages | 14 (100%) | 0 | 1 QRDR: 14 (100%) | 0 | 0 |

Acquired resistance genes were almost exclusively integrated into the *S.* Typhi chromosome, most frequently mediated by the translocation of a MDR Tn*2670*-like composite transposon (carrying genes *catA1, dfrA7, blaTEM-1, strAB, sul1* and *sul2*; 38.4% of all genomes in the main surveillance periods), and occasionally by transposon Tn*2670* (carrying genes *catA1, dfrA7,* and *sul1*; n=27) or Tn*6029* (carrying genes *blaTEM-1, strAB, sul2*; n=1). The exception to this was the *qnrS* gene, which was carried by an IncFIBK plasmid (also carrying genes *blaTEM-1, sul2, tet(A)*, n=12). In Dhaka, MDR *S*. Typhi was significantly less common in older age groups (OR 0.70, [95% CI, 0.52-0.94], p=0.02) (**Fig. S1b**).

Antimicrobial susceptibility phenotypes were available for a subset of isolates and non-susceptibility was mostly well-predicted by known molecular mechanisms (in *S.* Typhi, 99% positive predictive value for first-line drugs and >96% for ciprofloxacin; in *S.* Paratyphi A, 99% positive predictive value for ciprofloxacin; **Tables S4-5**). Phenotypic assessment of azithromycin susceptibility is challenging^23^ and susceptibility thresholds are poorly defined. Three of five *acrB* mutants tested resistant for azithromycin. Several other isolates showed azithromycin-resistant phenotypes but had wildtype *acrB* genes and 23S alleles, no acquired macrolide resistance genes, and a genome-wide screen did not identify any novel variants associated with the reported phenotypes (see **Supplementary Methods**).

We used ancestral state reconstruction of AMR determinants on a global phylogeny to differentiate emergence of AMR from transmission of resistant strains (summarised in **Table 3**). In Blantyre, all MDR cases were attributed to local transmission of MDR *S*. Typhi 4.3.1.1.EA1 (**Fig. S4**)^9^. In contrast, all three instances of QRDR mutations in Blantyre were attributed to local evolution, arising independently in the endemic MDR *S*. Typhi 4.3.1.1.EA1 strain background and with no evidence of transmission to secondary cases (**Fig. S4**).

**Table 3.** AMR transmission. Each row represents a specific AMR pattern identified in a given serovar (Ty=Typhi, PA=Paratyphi A) at a given site, and summarises the number of cases sequenced; the number of independent lineages those cases cluster into in the phylogeny; and the number of resistant cases attributed to transmission of (i) local or (ii) imported resistant variants, or (iii) *de novo* evolution of resistance. These three categories (i-iii) were assigned based on ancestral state reconstruction of AMR determinants and country on the global-context phylogenies, full details and definitions are given in Supplementary Methods. ‘ARG’, antimicrobial resistance gene; ‘QRDR’, quinolone resistance determining region; ‘1 QRDR’ indicates one such mutation (associated with decreased ciprofloxacin susceptibility); ‘3 QRDR’ indicates three such mutations (associated with ciprofloxacin resistance); AcrB indicates *acrB* mutation associated with azithromycin resistance.

|  | Serovar | Resistant Cases<br>N (% of serovar) | Clusters<br>N | Source of resistance, N (% of resistant) |  |  |
| --- | --- | --- | --- | --- | --- | --- |
|  |  |  |  | (i) Transmitted,<br>local variant | (ii) Transmitted,<br>imported variant | (iii) *De novo*<br>evolved variant |
| Blantyre |  |  |  |  |  |  |
| First-line drug ARGs | Ty | 138 (97.9%) | 1 | 138 (100%) | 0 | 0 |
| 1 QRDR | Ty | 3 (2.1%) | 3 | 0 | 0 | 3 (100%) |
| Dhaka |  |  |  |  |  |  |
| First-line drug ARGs | Ty | 170 (47.6%) | 2 | 170 (100%) | 0 | 0 |
| 1 QRDR | Ty | 343 (96.1%) | 13 | 343 (100%) | 0 | 0 |
|  | PA | 95 (100%) | 4 | 94 (99%) | 0 | 1 (1%) |
| 3 QRDR | Ty | 1 (0.3%) | 1 | 0 | 1 (100%) | 0 |
| QRDR + *qnr* | Ty | 12 (3.4%) | 1 | 12 (100%) | 0 | 0 |
| AcrB-R717Q | Ty | 2 (0.6%) | 2 | 1 (50%) | 0 | 1 (50%) |
|  | PA | 3 (3.2%) | 2 | 2 (66%) | 0 | 1 (33%) |
| Kathmandu |  |  |  |  |  |  |
| First-line drug ARGs | Ty | 2 (1.6%) | 2 | 2 (100%) | 0 | 0 |
| 1 QRDR | Ty | 106 (85.5%) | 8 | 103 (97.2%) | 1 (0.9%) | 2 (1.9%) |
|  | PA | 14 (100%) | 3 | 13 (93%) | 1 (7%) | 0 |
| 3 QRDR | Ty | 3 (2.4%) | 1 | 3 (100%) | 0 | 0 |

In Dhaka, all resistance to first-line drugs was attributed to local transmission of *S*. Typhi 4.3.1.1 (MDR) and 4.3.1.3.Bdq (ampicillin-resistant) populations that had become endemic prior to the surveillance period (**Fig. S5**)^2, 33^. Nearly all of the 451 sequences carrying QRDR mutations in Dhaka (n=356 *S.* Typhi, n=95 *S.* Paratyphi A) were also attributed to local transmission of endemic strains (**Table 3, Fig. S5**). The exceptions were one *S*. Paratyphi A 2.4.4 (**Fig. S3c; de novo evolution**), and one *S*. Typhi 4.3.1.2 that was likely imported from India^3, 7, 10^ (**Fig. S3d**). Notably, the other ciprofloxacin-resistant *S*. Typhi cases in Dhaka were all attributed to local transmission of the endemic strain 4.3.1.3.Bdq (**Fig. S5**). AcrB-R717Q mutations were either inherited from locally established populations (n=1 *S.* Typhi 4.3.1.1, **Fig. S3e**; n=2 *S.* Paratyphi A 2.4.4, **Fig. S3c**), or arose independently in local populations (n=1 *S.* Typhi 3.3.2, **Fig. S3f**; n=1 *S.* Paratyphi A 2.4.4, **Fig. S3c**).

In Kathmandu, resistance to former first-line drugs was rare (n=2, 1.4%), occurred only in *S.* Typhi 4.3.1.1 (**Fig. S6**), and resulted from local transmission of endemic strains pre-dating STRATAA surveillance (**Table 3**). Most cases (85.5% of *S.* Typhi, all *S.* Paratyphi A) carried a single QRDR mutation. These result from 11 different clusters, each with a characteristic *gyrA* mutation (see **Fig. S6**), most of which (97% of *S*. Typhi, 93% of *S*. Paratyphi A) were already present in the local population and carrying QRDR mutations before the surveillance period (**Table 3**). The exceptions were one *S.* Paratyphi A 2.3 (Fig. S3g) and one *S*. Typhi 4.3.1.1 (Fig. S3h) that were closely related to sequences from India (8-9 single nucleotide variants (SNVs)); and two*S.* Typhi 3.2.2 with no close relatives in the genome collections (Fig. S3a).

### Transmission dynamics

Substitution mutations accumulate too slowly in *S.* Typhi and *S.* Paratyphi A to infer specific transmission events from sequence data^24^; hence, to explore transmission patterns we instead examined groups of cases in the STRATAA dataset that formed zero-SNV clusters (i.e. no SNVs detected in the core genome), which we interpret as being linked either by a common source or by chains of transmission during which no substitutions have arisen. Two-thirds (67%) of cases fell into zero-SNV clusters (60% in Blantyre, 69% in Dhaka, 59% in Kathmandu). Most clusters were of size n=2 cases (40 clusters) or n=3 cases (22 clusters), but there were also 25 large clusters of ≥5 cases (accounting for 21% of cases in Blantyre, 51% in Dhaka, 30% in Kathmandu). The median time between consecutive cases in the same cluster was 16 days (interquartile range 3-49 days). Median time between first and last cases per cluster was 136 days (interquartile range 37–273 days, i.e. ∼1.2–9.1 months) and the maximum was 854 days (28 months, i.e. 2.3 years); zero-SNV clusters spanning >1 year were detected in all sites. This confirms the slow substitution rate and highlights the difficulty in resolving individual transmission events for typhoidal pathogens. Cases from the same zero-SNV clusters showed significant temporal clustering (reduced pairwise temporal distances compared to unclustered isolates; see **Fig. 3**) in all settings and for both serovars.

**Figure 3:**
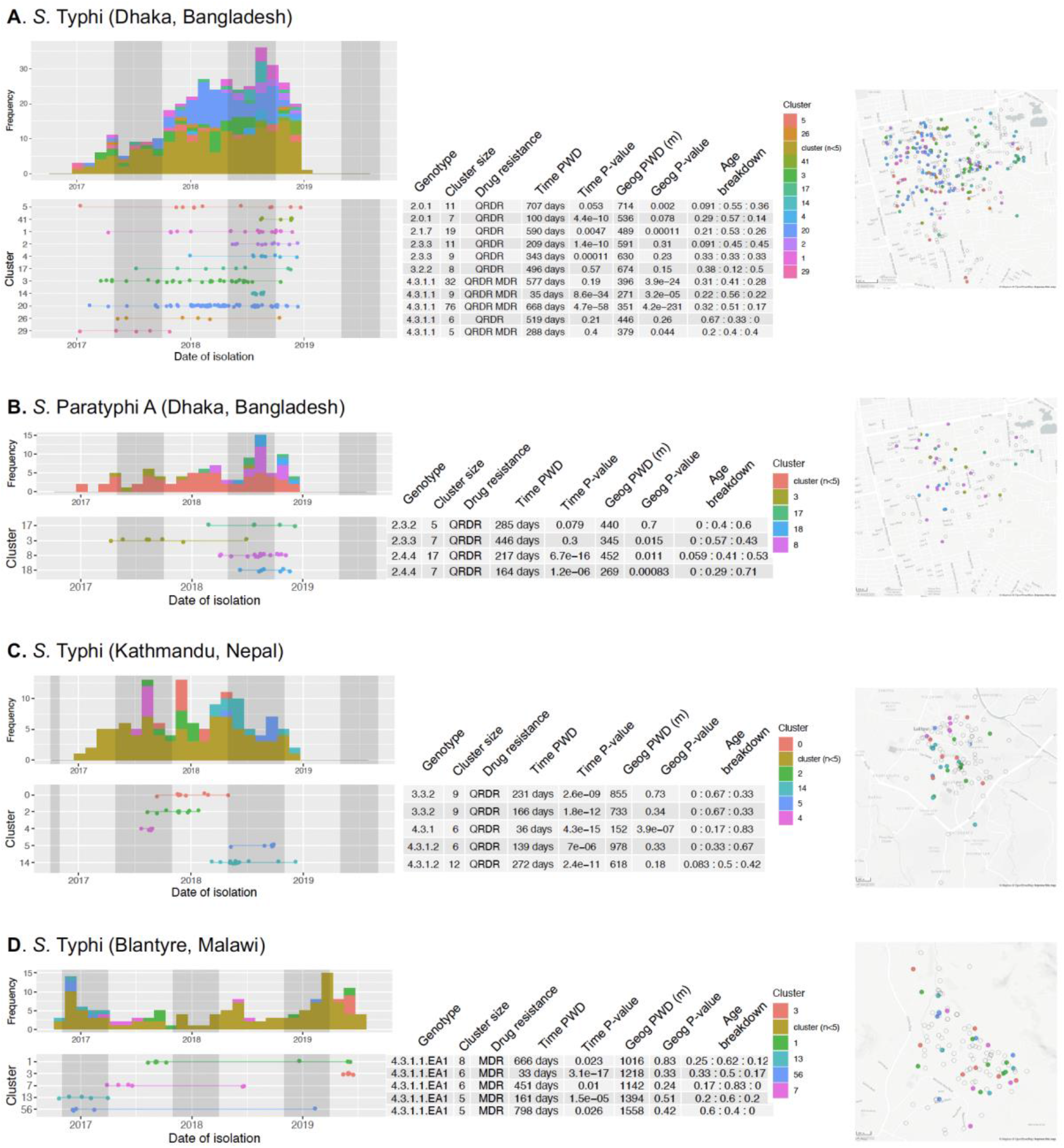
Features of zero-SNV clusters. Each panel (A-D) summarises clusters for a given city and serovar, as indicated; data are shown for common zero-SNV clusters (comprising at least five cases). Within each panel, from left to right is shown: monthly counts for each cluster above a time-line (one row per cluster, as labelled); table summarising key information per cluster (rows as per timeline figure); legend indicating colour code per cluster, which applies across the whole panel; and spatial distribution of clusters on a map of the catchment area. In the table, ‘Time’ and ‘Geog’ p-values are the result of Kolmogorov-Smirnov tests comparing the pairwise distribution within a given cluster to the distribution between all unclustered isolates from that site. Darker shaded grey vertical areas represent the rainy season in each location. Filled points on the map each represent a sequenced case, coloured by the zero-SNV cluster as per inset legend in each panel (unfilled points indicate other cases, including unclustered cases or members of smaller clusters).

There was some evidence of geographical clustering of zero-SNV cases compared to unclustered cases in the South Asian settings (*S*. Typhi and *S*. Paratyphi A in Dhaka, median distance 373 m vs 540 m and 421 m vs 524 m, respectively; *S*. Typhi in Kathmandu, 646 m vs 796 m). **Fig. 3** shows properties of the 25 larger zero-SNV clusters (≥5 cases). Just 10 of these showed evidence of spatial clustering, as might be expected for transmission following a point-source outbreak: nine in Dhaka (*S*. Typhi 2.0.1, 2.1.7, and 4.3.1.1, spanning up to ∼2 years each; *S*. Paratyphi A 2.3.3 and 2.4.4, ∼14 months) and one in Kathmandu (*S*. Typhi 4.3.1, 36 days) (see **Fig. 3**).

AMR profiles were homogeneous within clusters (**Fig. 3**), consistent with transmission of AMR variants. In Kathmandu, cases caused by the largely imported strain *S*. Typhi 3.3.2 were more likely to be in zero-SNV clusters (71%, in six clusters) compared with the endemic *S*. Typhi 4.3.1.2 (61%, 11 clusters), other *S*. Typhi (38%, 1 cluster) or *S*. Paratyphi A (36%, 2 clusters) (p=0.03 using Chi-square test). In Dhaka, some genotypes had significantly higher clustering rates (4.3.1.1, 82%; 4.3.1.3, 75%; 2.0.1, 68%) than others (2.3.3, 59%; *S*. Paratyphi A, 62%) (p<1x10^-6^ using Chi-square test), which may reflect greater intensity of transmission^20^. In all settings, all three age groups were equally likely to be involved in zero- SNV clusters (64-66% per age group, p>0.4 in all settings, using Chi-square test), and most clusters (77%) were detected across age groups (**Fig. 3**).

## Discussion

This study provides a comprehensive, WGS-resolved, view of the pathogen populations underlying enteric fever in three urban sites in areas of high disease burden. Use of the same passive surveillance protocols across all sites, in defined study catchments and including all ages, allows the results to be compared between sites. The genotype prevalences we estimated are in line with other studies of enteric fever pathogens reported from the same cities^2, 3, 7–10, 21, 25, 26^ , but we also specifically addressed pathogen diversity across age groups. We show that although the disease burden is highest in children^20^, the same pathogen variants (serovars, genotypes, and AMR) circulate across age groups (pre-school, school-age, and adult), with near-identical sequences (0-SNV clusters) identified across age groups (**Figs. 2- 3, S4-S6**). This supports the assumption of homogeneous mixing between age groups in models of TCV impact. It also supports the idea that vaccinating children may have a secondary impact by reducing transmission to adults, although the directionality of transmission is not resolvable from WGS and no statistically significant effect has yet been observed in TCV trials.

Importantly, this study explicitly addresses the transmission burden of drug-resistance in enteric fever (**Table 3**). Several prior studies have documented transmission of AMR variants between countries, including introduction of MDR 4.3.1 variants from South Asia into Kenya, Malawi and neighbouring countries^8, 9, 27^, and spread of ciprofloxacin-resistant 4.3.1.2 from India into Nepal^3, 10^. A recent study estimated QRDR mutations have arisen ≥94 times in *S*. Typhi and transferred between countries ≥119 times^7^. It is also frequently reported that AMR infections result from clonal spread. However, whilst several studies have used WGS and ancestral state reconstruction to estimate the proportion of AMR *Mycobacterium tuberculosis* infections that are due to transmission of AMR variants vs *de novo* emergence^28^, this has not been previously quantified in enteric fever pathogens. Our results show that the vast majority of enteric fever in all three study sites was due to circulation of locally-established pathogen variants, and that AMR infections were overwhelmingly (98%) caused by transmission of pre-existing AMR strains (**Table 3**). This supports the modelling assumption that each AMR infection poses a risk of secondary AMR infections in others, through shedding and onward transmission of the AMR variants. Coupled with our data showing the same variants co-circulate freely across age groups, this lends weight to model predictions that reducing the incidence of AMR *S*. Typhi infection and shedding in children through TCVs may have a secondary effect of reducing exposure to AMR infections across the whole population.

Our data showing persistence of zero-SNV clusters over months and even years (**Fig. 3**) highlights the challenges of inferring transmission events from *S*. Typhi WGS data. Previous studies have estimated substitution rates of approximately one SNV every two years^3, 8^, a log- scale slower than for host-generalist *S. enterica* serovars such as Agona and Kentucky, and similar to slow-growing *M. tuberculosis*^29^. Our data support previous assertions that the evolutionary rate in *S*. Typhi is too slow to support inference of explicit transmission events and networks from genome sequence data^24^; although it is still possible to obtain useful data on transmission dynamics from WGS data (such as estimates of R0 and trends in effective population size^7^). Simple spatial analysis using GPS coordinates without topological mapping identified significant spatial clustering in Dhaka and Kathmandu but not Blantyre (**Fig. 3**).

The Blantyre study area has a complex river system, comprising 10 river catchments in close proximity, and recent spatial modelling of *S*. Typhi sequences in this setting showed that dividing space into river catchments explained the spatio-genetic patterns much better than simple grid coordinates^26^. Detailed spatial modelling is beyond the scope of this study but is planned for all STRATAA sites.

Overall, our findings strengthen efforts to model the impact of TCVs, and provide essential baseline data against which to assess the impact of future immunisation programmes and other interventions to control enteric fever.

## Methods

### Ethics statement

Research Ethics Committee approval for a joint study protocol across all three surveillance sites was obtained within each country as well as from the Oxford Tropical Research Ethics committee as detailed elsewhere^20, 22^.

### Study protocol

The STRATAA study protocol has been previously published^22^, and subsequent findings described in detail elsewhere^23^. Antimicrobial susceptibility testing was done using disc diffusion^23^.

### Whole genome sequencing and analysis

Blood culture isolates were subjected to WGS via Illumina HiSeq to generate 100 bp paired-end reads as described previously^8^. SNVs were identified by mapping sequence reads to the *S*. Typhi CT18 (accession no. AL513382) and *S*. Paratyphi A AKU_12601 (accession no. FM200053) reference genomes using RedDog (V1beta.11). Genotypes were assigned using GenoTyphi^30^ (v1.9.1) and Paratype. Recombination-filtered maximum-likelihood phylogenies were inferred using Gubbins (v2.4.1) and RAxML (v8.2.8). Acquired AMR genes and plasmid replicons were detected using SRST2 (v0.2.0). To determine if detected molecular determinants of AMR were transmitted or inherited, we performed maximum-parsimony ancestral state reconstruction of AMR determinants and country, using R package *phangorn* (v2.5.5). See **Supplementary Methods** for full details of sequencing and analysis.

### Statistical analyses

Statistical tests were conducted in R (v4.1.2) using two-sided tests of significance. Logistic regression models were fit using the *glm*() function in package *stats* (v4.1.2). Age-group was treated as an ordinal categorical variable (<5 years, ≥5 and <15 years, ≥15 years). Severe disease was defined as symptom duration exceeding 10 days and/or requiring hospitalisation (based on clinical review of case report forms). Further details are provided in **Supplementary Methods**.

### Nucleotide sequence data accession numbers

Raw sequence data have been deposited in the European Nucleotide Archive under project PRJEB14050 (accessions in **Table S1**).

## Data Availability

Raw sequence data have been deposited in the European Nucleotide Archive under project PRJEB14050 (accessions in Table S1). All other data produced in the present work are contained in the manuscript.

## Acknowledgements

This research was funded by the Wellcome Trust [STRATAA grant number 106158/Z/14/Z and core funding to the Wellcome Sanger Institute, grant number 206194] and the Bill & Melinda Gates Foundation [grant number OPP1141321]. ZAD received funding from the European Union’s Horizon 2020 research and innovation programme under the Marie Skłodowska-Curie grant agreement No 845681. For the purpose of open access, the author has applied a CC BY public copyright licence to any Author Accepted Manuscript version arising from this submission. We acknowledge the contribution of all participants who have taken part in the studies and the large field and laboratory teams at the sites, including: Amit Aryja, Binod Lal Bajracharya, David Banda, Yama Mujadidi, Pallavi Gurung, Nazia Rahman, Archana Maharjan, George Mangulenji, Prasanta Kumar Biswas, and the Nepal Family Development Foundation team. We also acknowledge the sequencing and pathogen informatics teams at the Wellcome Sanger Institute for sequencing and data processing; and Sebastian Duchene of the University of Melbourne for useful discussions regarding transmission analyses. icddr,b is grateful to the Government of Bangladesh, Canada, Sweden and the United Kingdom for support.

## Contributors

AJP, BB, FQ, GD, JDC, KEH, MG, RSH, SB, SD, VEP designed the study and obtained funding. They oversaw data collection together with AK, CM, NF, MS. AC, AK, FK, MS, JM contributed to data collection and curation. ST was project manager. ZAD, KEH, PA analysed data; AJP, JM, MG, RSH, SB, VEP assisted with design and interpretation of the analysis. ZAD and KEH had full access to all the data in the study and verified the data. ZAD, KEH and PA wrote the first draft of the manuscript, and all authors reviewed the manuscript critically for content and approved the decision to submit for publication.

## STRATAA Study Group Authorship

Anup Adhikari^5^, Happy Chimphako Banda^6^, Christoph Blohmke^1^, Thomas C. Darton^1^, Christiane Dolecek^2^, Yama Farooq^1^, Maheshwar Ghimire^5^, Jennifer Hill^1^, Nhu Tran Hoang^3^, Tikhala Makhaza Jere^6^, Moses Kamzati^6^, Yu-Han Kao^4^, Clemens Masesa^6^, Maurice Mbewe^6^, Harrison Msuku^6^, Patrick Munthali^6^, Tran Vu Thieu Nga^3^, Rose Nkhata^6^, Neil J. Saad^4^, Trinh Van Tan^3^, Deus Thindwa^6^, Merryn Voysey^1^

## Affiliations

^1^ Oxford Vaccine Group, Department of Paediatrics, University of Oxford, and the NIHR Oxford Biomedical Research Centre, Oxford, United Kingdom

^2^ Nuffield Department of Medicine, Centre for Tropical Medicine and Global Health, University of Oxford, Oxford, UK

^3^ The Hospital for Tropical Diseases, Wellcome Trust Major Overseas Programme, Oxford University Clinical Research Unit, Ho Chi Minh City, Vietnam

^4^ Department of Epidemiology of Microbial Diseases and the Public Health Modeling Unit, Yale School of Public Health, Yale University, New Haven, CT, USA

^5^ Oxford University Clinical Research Unit, Patan Academy of Health Sciences, Kathmandu, Nepal

^6^ Malawi-Liverpool Wellcome Programme, Blantyre, Malawi

## Supplementary Methods

### Bacterial isolates and whole genome sequencing

Isolates cultured from blood of febrile individuals recruited into the STRATAA passive surveillance studies at each of the three sites were stored locally until the end of the recruitment period. Subsequently, isolates were cultured overnight and genomic DNA extracted using the Wizard Genomic DNA Extraction Kit following the manufacturers recommendations (Promega, WI, USA). DNA was shipped to the Wellcome Sanger Institute and subjected to indexed whole genome sequencing on an Illumina HiSeq 2500 HiSeq platform to generate paired-end reads of 100 bp in length, as described previously^1^.

Isolates from n=452 cases in Dhaka (99.6% of all culture-positive) from the period January 2017-December 2018 were successfully sequenced and confirmed as *S*. Typhi or *S*. Paratyphi A (summary in **Table 1,** genome list in **Table S1**). From Kathmandu, n=138 cases (84.1% of all culture-positive) from the same time period were successfully sequenced and serovars confirmed. From Blantyre, n=83 cases (72.2% of all culture-positive) from October 2016-October 2018 were successfully sequenced and typhoidal serovars confirmed. Recruitment continued for an additional 10 months in Blantyre, resulting in total n=141 sequenced typhoidal isolates (89.2% of positive cultures). The full set of n=141 sequences from Blantyre are included in the main dataset ‘STRATAA’, which comprises genome sequence data for n=731 unique typhoidal *Salmonella* blood-culture isolates (622 *S*. Typhi and 109 *S*. Paratyphi A, see **Table 1**). A small number of additional blood-culture isolates were captured at the study sites before or after these formal surveillance periods (n=30 from Dhaka, n=27 from Kathmandu) or outside the boundaries of the surveillance catchment area (n=35 from Blantyre); these were also sequenced and included in phylogenetic trees to provide context (total n=707 *S*. Typhi, n=116 *S*. Paratyphi A; see **Table S1**), but were excluded from statistical analyses.

### Single nucleotide variant (SNV) analysis and *in silico* genotyping

Raw *S.* Typhi Illumina reads were mapped to the *S.* Typhi CT18 reference sequence (accession no. AL513382)^2^, and those for *S.* Paratyphi A to the *S*. ParatyphiA AKU_12601 reference genome (accession no. FM200053)^3^.

Mapping was carried out using the RedDog mapping pipeline (V1beta.11; available at https://github.com/katholt/reddog), which uses Bowtie (v2.2.9)^4^ to map reads to the reference sequence, and SAMtools (v1.3.1)^5^ to identify single nucleotide variant (SNV) calls as previously described^6^. All raw read data analysed had a minimum read depth of >40-fold, and >97% reference coverage. For *S.* Typhi sequences, read alignments (BAM files) were then used as input for GenoTyphi (v1.9.1; available at: https://github.com/katholt/genotyphi)7 to assign *S.* Typhi isolates to known genotypes according to an extended *S.* Typhi genotyping framework^7, 8^. Similarly, Paratype (v1.0; available at https://github.com/CHRF-Genomics/Paratype) was used to assign *S*. Paratyphi A sequences to genotypes ^9^.

Chromosomal SNVs with confident homozygous base calls (phred score >20), for all SNV sites that had such calls in >95% of *S.* Typhi genomes (representing the 95% ‘soft’ core genome) were concatenated to form an alignment of alleles at 15,387 variant sites for all *S.* Typhi, including 707 from this study (**Table S1**) and 3,128 global context sequences (**Table S6**) from previous studies ^6, 10–19^. Previously defined^8, 20, 21^ repetitive and recombinant regions were excluded (354 kb; ∼7.4% of bases in the CT18 reference chromosome), and any remaining recombination filtered out using Gubbins (v2.4.1)^22^ resulting in a final alignment length of 14,780 chromosomal SNVs. *S.* Typhi phylogenies were outgroup rooted using *S.* Paratyphi A AKU_12601 alleles, and a representative selection of *S.* Typhi outgroup taxa from each defined genotype (**Table S7**). An interactive version of the *S*. Typhi phylogeny with STRATAA plus global isolates (shown in **Figure S2**) is available at https://microreact.org/project/wim5TssQ3AqSfgWXTSP2Bd. Genotype-specific phylogenies were constructed in the same manner (used for ancestral state reconstruction, described below); as were local site-specific phylogenies of STRAATA data (**Figures S4-S6**), and *S.* Paratyphi A phylogenies (outgroup-rooted using *S.* Typhi CT18 alleles, https://microreact.org/project/SgHbIR9cP).

### Phylogenomic analysis

Maximum-likelihood (ML) phylogenetic trees were inferred from the aforementioned chromosomal SNV alignments using RAxML (v8.2.8)^23^. A generalised time-reversible model and a Gamma distribution was used to model site-specific rate variation (GTR+Γ substitution model; GTRGAMMA in RAxML) with 100 bootstrap pseudoreplicates used to assess branch support for the ML phylogeny. The resulting phylogenies were visualised and annotated using Microreact ^24^ and the R package *ggtree* (v2.2.4)^25^.

### Identification of antimicrobial resistance (AMR) determinants and associated mobile genetic elements

For the detection of chromosomal point mutations associated with AMR, GenoTyphi (v1.9.1; available at: https://github.com/katholt/genotyphi)7,8 and GenoParatyphi (v0.1-alpha; available at: https://github.com/zadyson/genoparatyphi) were used. These tools screen for mutations in the quinolone resistance determining region (QRDR) of *gyrA* (codons 83 and 87), *parC* (codons 80 and 84) and *gyrB* (codon 464), and mutations at *acrB* codon 717 (azithromycin resistance)^26, 27^. Acquired genes and plasmid replicons were detected using the mapping-based allele typer SRST2 (v0.2.0)^28^ to screen against the ARG-ANNOT^29^ and PlasmidFinder^30^ databases, respectively.

### Comparison of AMR genotypes and phenotypes

To assess how well genetic determinants predict AMR phenotypes, we interpreted the following markers as predictive of non-susceptibility to specific antimicrobials, based on previous publications^27, 31^: *bla*_TEM-1_, ampicillin; carbapenemase, meropenem; *catA1*, chloramphenicol; *dfr* plus *sul*, co-trimoxazole; extended-spectrum beta-lactamase, ceftriaxone and cefixime; *acrB*-717 mutation or *mphA*, azithromycin; QRDR mutation or *qnrS*, ciprofloxacin and nalidixic acid. Results are reported for drugs that were tested in ≥20% of isolates.

As most azithromycin resistance was not explained by known mechanisms (*acrB*-717 mutation or acquired genes), we screened Ribosomal RNA (rRNA) operons for potential explanatory mutations. A single copy of the rRNA operon (coordinates 4257263-4262892 in the CT18 reference genome) was extracted, and used as a reference sequence for mapping of *S.* Typhi reads using RedDog (as described above). Read alignments (BAM format) were subjected to low-frequency variant calling LoFreq (v2.1.3.1)^32^, and SNV read-depth data extracted from the resultant variant calling format (VCF) files with the *read.vcfR*() function from the R package *vcfR* (v1.12.0)^33^. Read depths for SNVs detected in the rRNA operon were normalised using the average chromosomal read depths for each sequence reported by RedDog across the entire CT18 reference sequence to identify single copy mutations (read depth of ∼1x the average reported for the CT18 chromosome). These single copy mutations were examined for an association with observed azithromycin resistance phenotypes, but were found to correlate with the genotype backgrounds in which they occurred and not with resistance. The same approach was used to assess *S*. Paratyphi A genomes, where reads were mapped to a single rRNA operon (coordinates 3870557-3876131 in the *S*. Paratyphi A AKU_12601 reference genome), but again no evidence of resistance-associated mutations in the rRNA operon were identified.

DBGWAS (v0.5.4)^34^ was utilised to carry out a bacterial genome-wide association study (GWAS) to screen for genetic loci and/or variants associated with the azithromycin resistance. For this, raw reads were assembled *de novo* with Unicycler (v0.4.7)^35^ and used as input to DBGWAS, along with azithromycin resistance status for each sequence. DBGWAS was run using default parameters and p-value <0.01 interpreted as a significant association; none were identified.

### Transmission of molecular determinants of AMR

To determine if resistant enteric fever cases resulted from infection with locally-circulating resistant strains, imported strains, or emerged *de novo*, we carried out ancestral state reconstruction (ASR) analyses. For each genotype where molecular determinants of AMR were detected, we inferred a ML phylogeny including all available STRATAA genomes plus global contextual sequences of the same genotype (phylogenetic inference methods as detailed above). We then conducted ASR, for each detected AMR determinant and country of origin (Nepal, Bangladesh, Malawi, or other), onto these ML tree topologies using a maximum-parsimony method implemented in the *ancestral.pars*() function in the R package *phangorn* (v2.5.5)^36^. The inferred states for each variable (country and AMR determinant) were extracted for each internal node of each tree and used to infer, for each resistant isolate (tree tip), whether the resistance was most likely: (i) inherited from a local resistant strain (parent node and tip share same AMR determinant and location, interpreted as local transmission of a resistant strain); (ii) inherited from an imported resistant strain (parent node and tip share the same AMR determinant but parent node is located in a different country, interpreted as transmission of an imported resistant strain); (iii) recently emerged *de novo* (parent node lacks the AMR determinant, interpreted as recent emergence of resistance rather than transmission of an established resistant strain).

### Cluster analysis

SNP distances were calculated using disty (v0.1.0, https://github.com/c2-d2/disty). Zero-SNV clusters were then identified using the AgglomerativeClustering function of the Python package scikit-learn (v1.0.2) i.e.

AgglomerativeClustering(linkage=’complete’, distance_threshold=0.1, n_clusters=None).fit_predict(df).

Global Position System (GPS) coordinates were collected by field workers from the homes of cases using either study tablets or GPS machines. If a GPS signal could not be obtained at a particular location due to density, then the closest location where a signal could be obtained was recorded instead. Coordinates were available for 88.6% of cases (94.5% in Dhaka, 77.5% in Kathmandu, 80.9% in Blantyre). Spatial pairwise distance was calculated using the geopy.distance.distance function from the geopy Python package (v2.3.0). Distributions of pairwise geographic and temporal distances were compared between cases in zero-SNV clusters and unclustered cases using the Kolmogorov-Smirnov two sample test implemented in the scipy (v1.7.1) package of Python i.e. scipy.stats.ks_2samp.

### Statistical analyses

Statistical tests were conducted in R (v4.1.2) using two-sided tests of significance. SNV distances were calculated from the SNV alignment (described above) using snp-dists (v0.7.0), available at (https://github.com/tseemann/snp-dists). Simpson’s index diversity estimates were calculated using the *diversity*() function in the R package *vegan* (v2.5.6)^37^. Phylogenetic signals for age-group were quantified using the function *multiPhylosignal*() in the R package *picante* (v1.8.2)^38^ to calculate *K* statistic^39^ values. Multivariate logistic regression models were fit using the *glm*() function in package *stats* (4.1.2). Age-group was treated as an ordinal categorical variable (<5 years, ≥5 and <15 years, ≥15 years); other variables (serovar, dominant genotype, MDR, severity) were coded as binary data.

## Supplementary Methods

### Supplementary Figures

**Figure S1.**
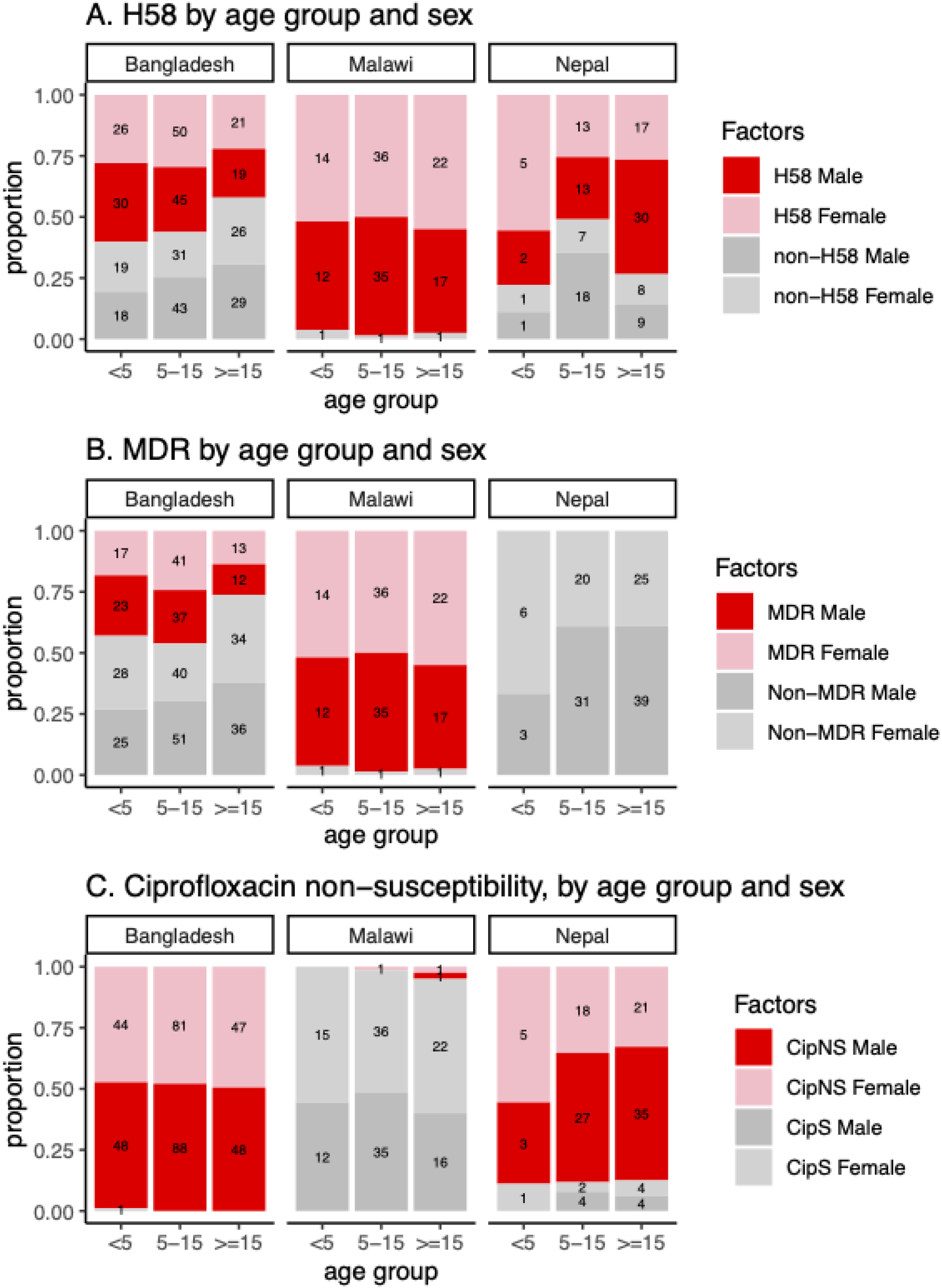
Breakdown of patient sex and *S*. Typhi features, within each age-group, at each site, for the main STRATAA dataset. (A) H58, genotype 4.3.1 and derived genotypes. **(B)** MDR, multi-drug resistant. **(C)** CipNS, ciprofloxacin non-susceptible; CipS, ciprofloxacin susceptible.

**Figure S2.**
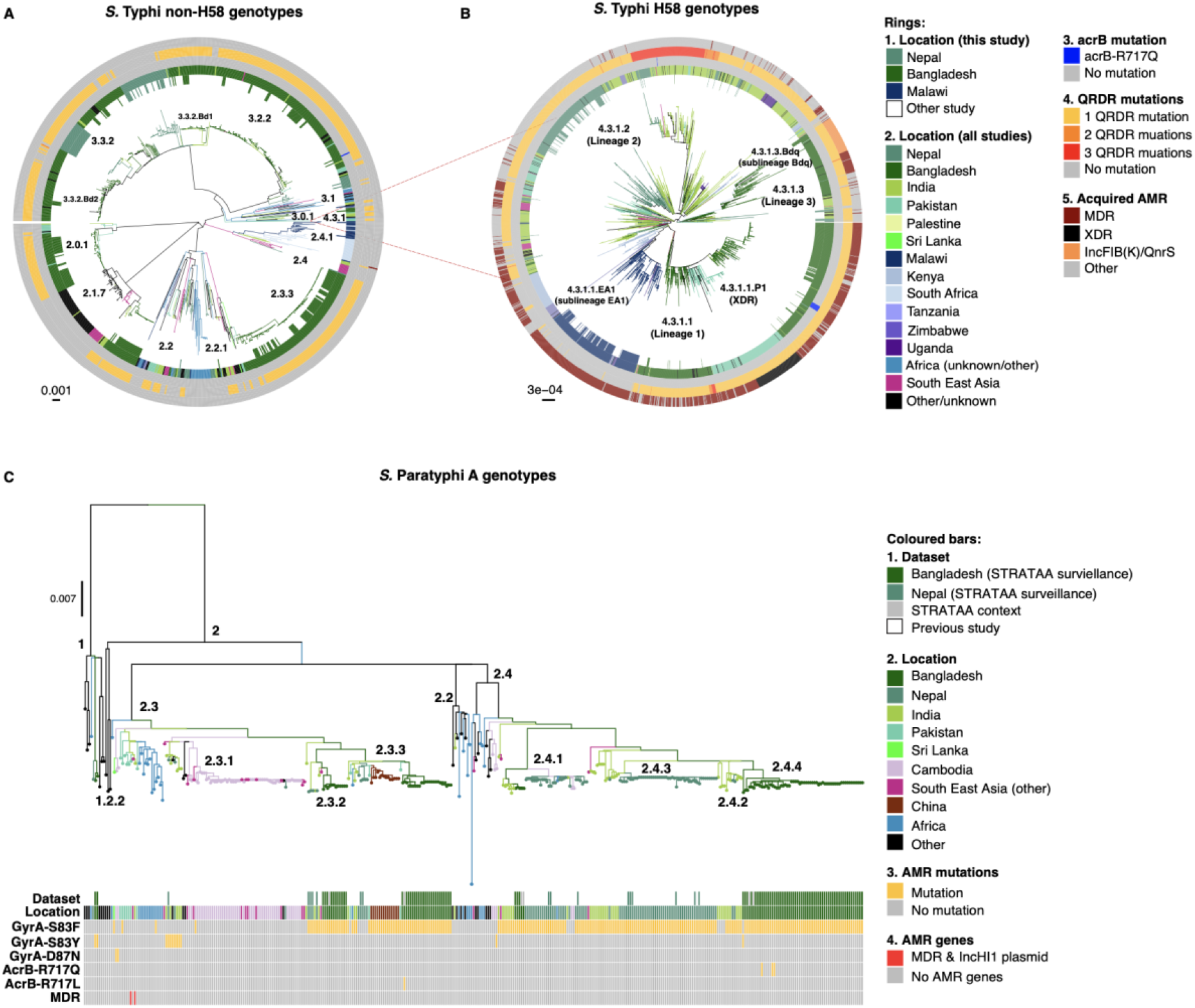
Phylogenetic trees of *S.* Typhi and S. Paratyphi A showing STRATAA isolate sequences in context with publicly available global genome data. (A) Non-H58 genomes (i.e., excluding 4.3.1 and derived genotypes). **(B)** Subtree of H58 (4.3.1 and derived) genotypes. A maximum-likelihood tree was inferred for n=3,835 *S.* Typhi sequences, including n=707 from this study plus global genomes, based on an alignment of genome-wide single nucleotide variants and outgroup rooted using *S.* Paratyphi A. The subtree of 4.3.1 genotypes (panel B) was pruned from the larger tree to aid visualisation. Genotypes are indicated with labels, and branch lengths indicate substitutions per variable site, as per inset scalebar. All branches and rings are coloured as per inset legend. Branch colours and first two rings indicate the geographic origin for the sequences; ring 1 highlights the n=707 sequences from this study. Other rings indicate antimicrobial resistance determinants as labelled. AcrB mutations are associated with azithromycin resistance. QRDR, quinolone resistance determining region. MDR, multidrug-resistant (MDR). XDR, extensively drug resistant. An interactive version of the full tree is available at https://microreact.org/project/wim5TssQ3AqSfgWXTSP2Bd.(C) **Paratyphi A genomes. A maximum likelihood phylogeny was inferred in the same manner as for (A) from n=375 genomes and outgroup rooted using S. Typhi. Branch colours and the first two coloured bars indicate geographic origin of the sequences; bar 1 highlights the n=116 sequences from this study. Other coloured bars indicate antimicrobial resistance determinants as labelled.** An interactive version of the full tree is available at: https://microreact.org/project/SgHbIR9cP.

**Figure S3.**
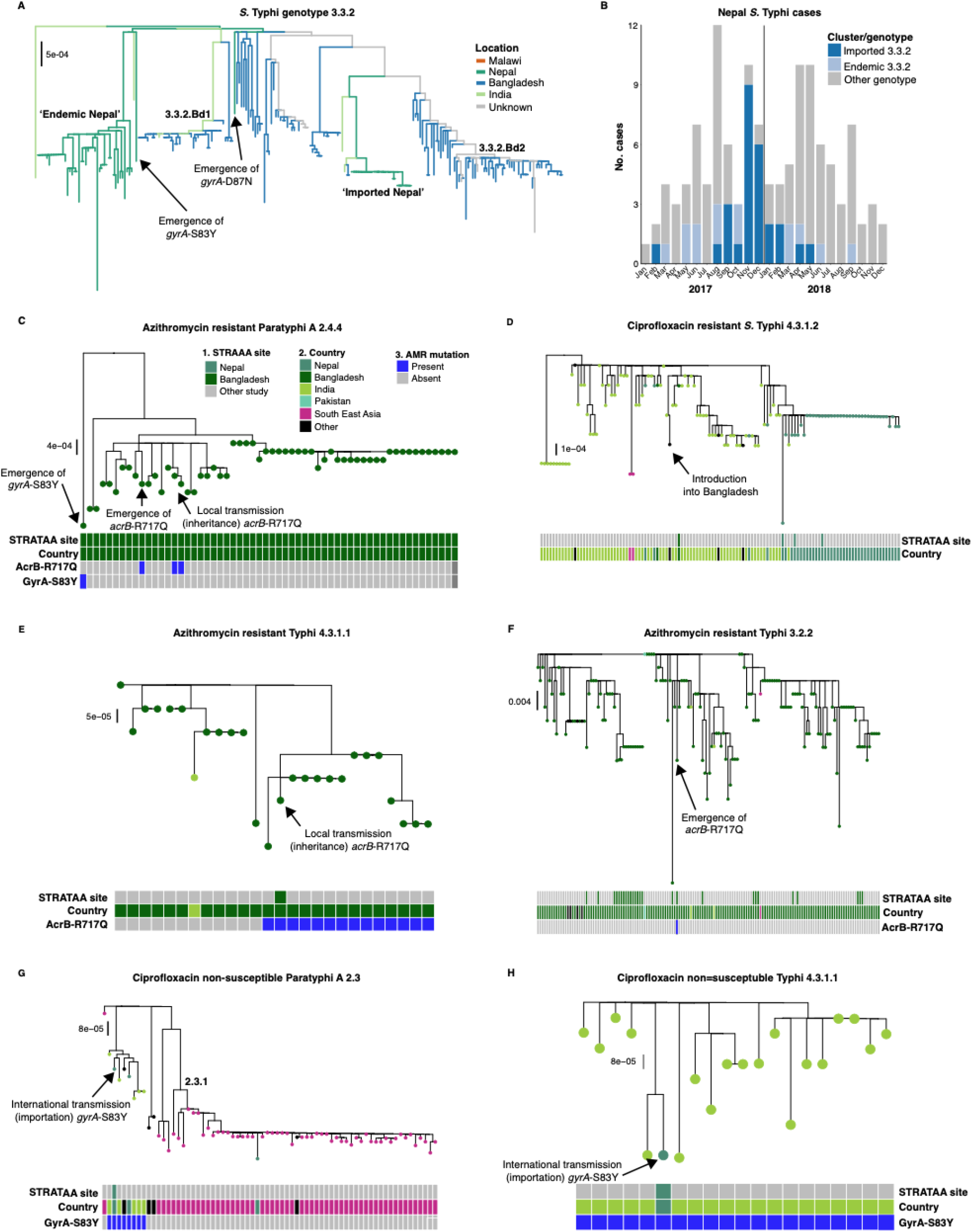
Detail of subpopulations discussed in text. Subtrees were extracted from the full tree of STRATAA plus global isolates (shown in **Figure S2**) to highlight specific points discussed in the manuscript. **(A)** *S.* Typhi 3.3.2 tree, highlighting the two distinct clades identified as being either endemic or imported in Nepal. Arrows indicate the de novo emergence of ciprofloxacin non-susceptibility driven by non-synonymous mutations in gyrA (as labelled). **(B)** Epidemic curve of *S.* Typhi sequenced from STRATAA surveillance in Kathmandu, showing counts of imported and endemic genotype 3.3.2, defined using the tree in panel A. **(C)** Tree highlighting the phylogenetic positions of three *acrB* mutant (i.e. azithromycin-resistant) *S*. Paratyphi A 2.4.4 sequenced from STRATAA surveillance in Dhaka (labelled with arrows). The population is mainly *acrB*- wildtype; two STRATAA isolates appear to share an *acrB* mutation inherited from a common ancestor, consistent with local transmission of an azithromycin-resistant variant; the third *acrB* mutant is distantly related from others and shows no evidence of transmission of a resistant ancestor. The emergence of the gyrA-S83Y mutation conferring ciprofloxacin non-susceptibility is also highlighted. **(D)** Tree highlighting the phylogenetic position of the single ciprofloxacin-resistant *S*. Typhi 4.3.1.2 sequenced from STRATAA surveillance in Dhaka (labelled with arrow), which suggests it was imported into Bangladesh (dark green) from India (light green) where it is believed to have emerged^10, 40^. **(E)** Tree highlighting the phylogenetic position of the single *acrB* mutant of *S*. Typhi 4.3.1.1 sequenced from STRATAA surveillance in Dhaka (labelled with arrow), which suggests it results from local transmission of an *acrB* mutant (i.e. azithromycin-resistant) clade that has been detected in previous studies from Dhaka^26^ (2013-2016). **(F)** Tree highlighting the phylogenetic position of the single *acrB* mutant (i.e. azithromycin-resistant) *S*. Typhi 3.2.2 sequenced from STRATAA surveillance in Dhaka (labelled with arrow); this suggests the mutation arose locally, in the background of a locally-circulating clade with wildtype *acrB. This* was categorised as a *de novo* resistance mutation. (G) Tree highlighting the phylogenetic position of the single gyrA-S83Y mutant (i.e. ciprofloxacin non-susceptible) S. Paratyphi A genotype 2.3 sequenced during STRATAA surveillance in Kathmandu (labelled with arrows). The position in the phylogeny suggests it was imported into Nepal (dark green) from India (light green). (H) Tree highlighting the phylogenetic position of the single gyrA-S83Y mutant (i.e. ciprofloxacin non-susceptible) S. Typhi genotype 4.3.1.1 sequenced during STRATAA surveillance in Kathmandu (labelled with arrows). The position in the phylogeny suggests it was imported into Nepal (dark green) from India (light green). For panels (**C-H**), branches are coloured by country of origin and coloured bars indicate samples from this study (STRATAA) by site, country of origin, and AcrB/GyrA status, as per the legend in panel (C).

**Figure S4.**
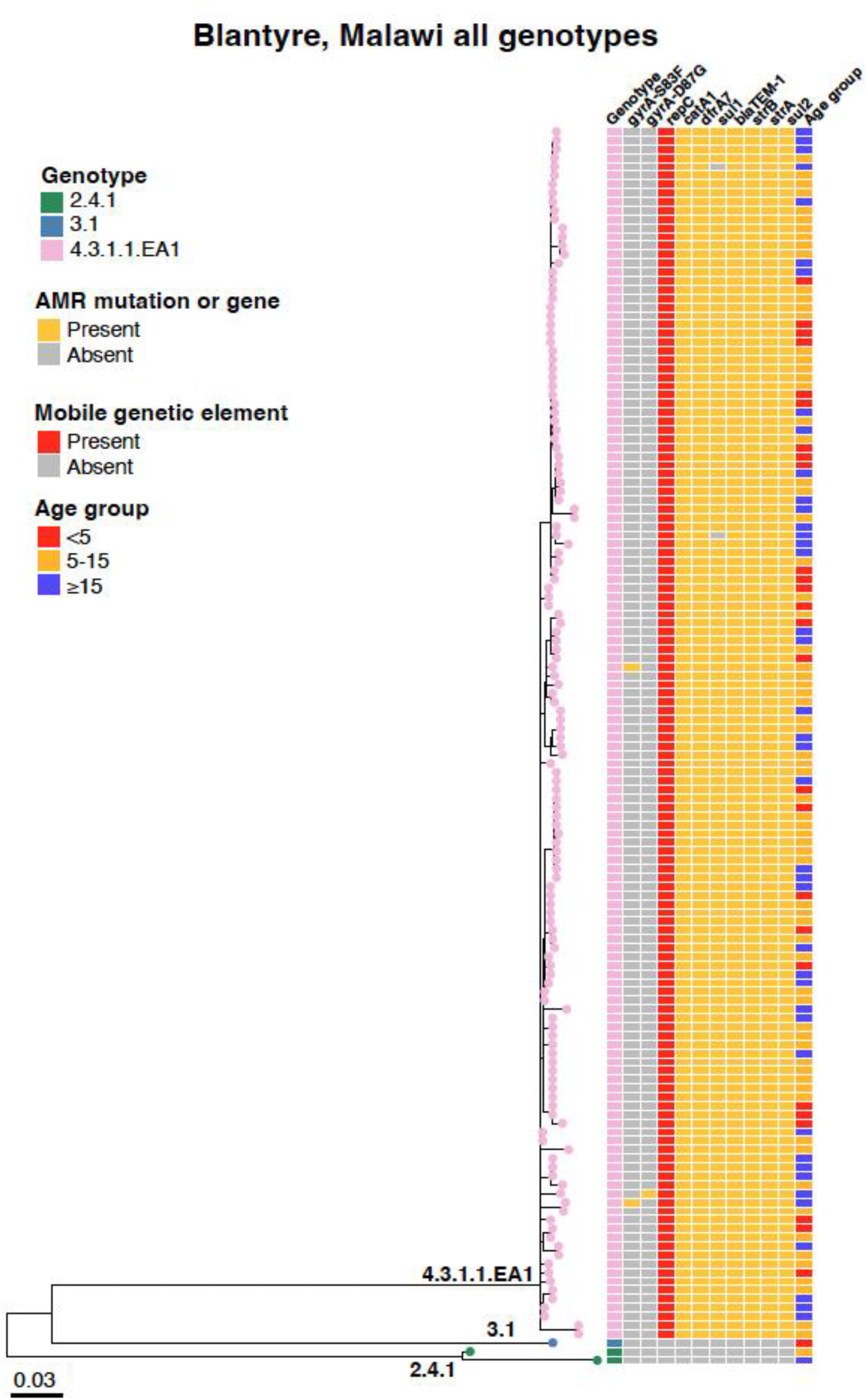
Phylogenetic tree of *S.* Typhi sequenced from STRATAA surveillance in Blantyre. Whole-genome maximum-likelihood phylogeny of n=141 *S.* Typhi sequences from Blantyre, Malawi (main ‘STRATAA’ dataset). Tip colours and first column of the heatmap indicate the genotype of the sequenced isolate (as per inset legend and branch labels). Second and third columns indicate presence of quinolone resistance-associated mutations in *gyrA*, remaining columns indicate presence of *repC* (a marker of the common multidrug resistant transposon), acquired genes associated with resistance to first-line drugs, and patient age group (coloured as per inset legends). Branch lengths indicate substitutions per variable site, as per the inset scalebar. Interactive phylogeny available at: https://microreact.org/project/beTX7esTof71d6J329iagM

**Figure S5.**
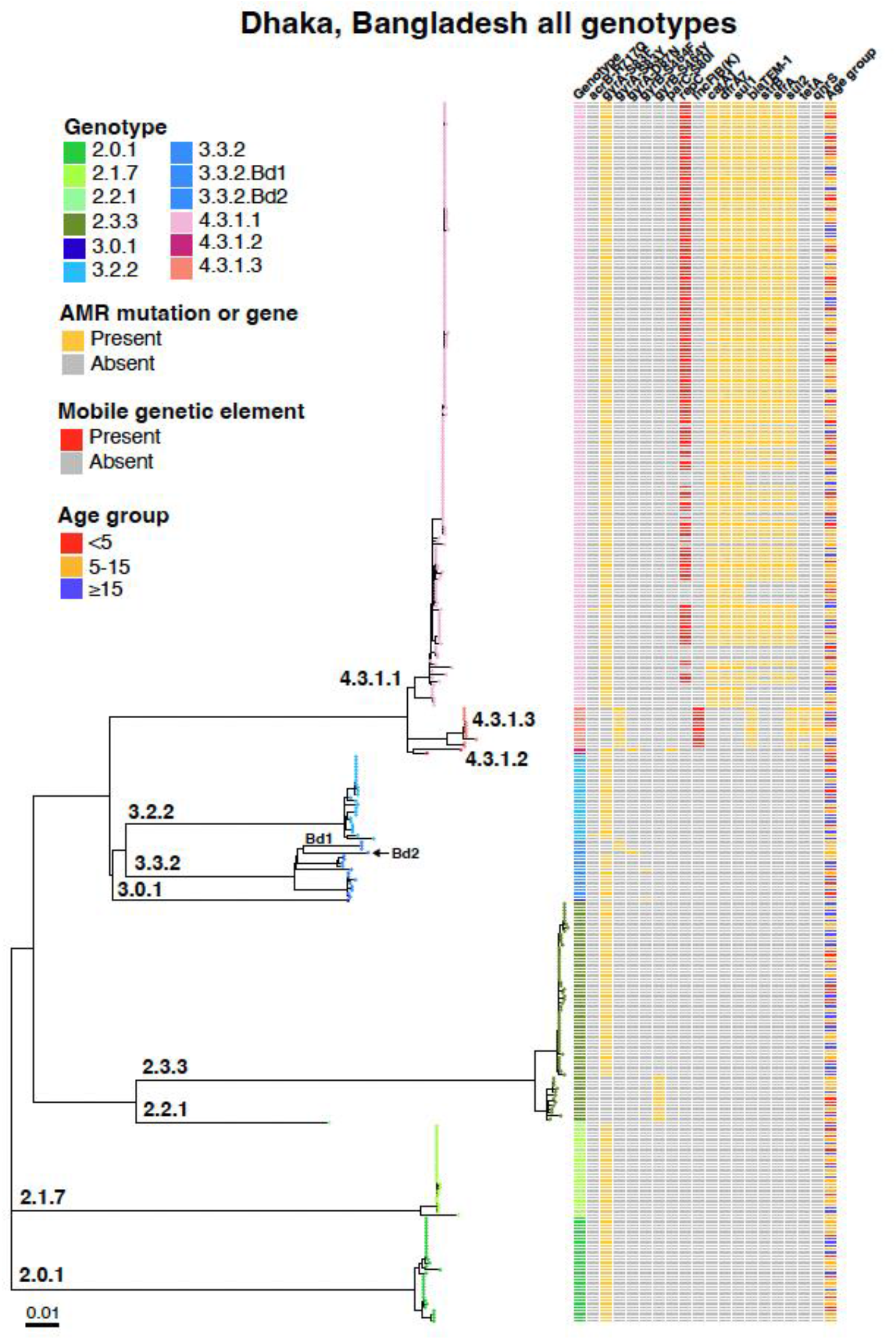
Phylogenetic tree of *S.* Typhi sequenced from STRATAA surveillance in Dhaka. Whole-genome maximum-likelihood phylogeny of n=357 *S.* Typhi sequences from Dhaka, Bangladesh (main ‘STRATAA’ dataset). Tip colours and first column of heatmap indicate the genotype of the sequenced isolate (as per inset legend and branch labels). Second column indicates presence of azithromycin resistance-associated mutations in gene *acrB*. Subsequent columns indicate the presence of quinolone resistance-associated mutations in *gyrA* and *parC*; *repC* (a marker of the common multidrug resistant transposon); the IncFIBk plasmid replicon marker; acquired drug resistance genes, and patient age group (coloured as per inset legends). Branch lengths indicate substitutions per variable site, as per inset scalebar. Interactive phylogeny available at: https://microreact.org/project/eAY3YZmVd6tXbWKijzZ5Fc.

**Figure S6.**
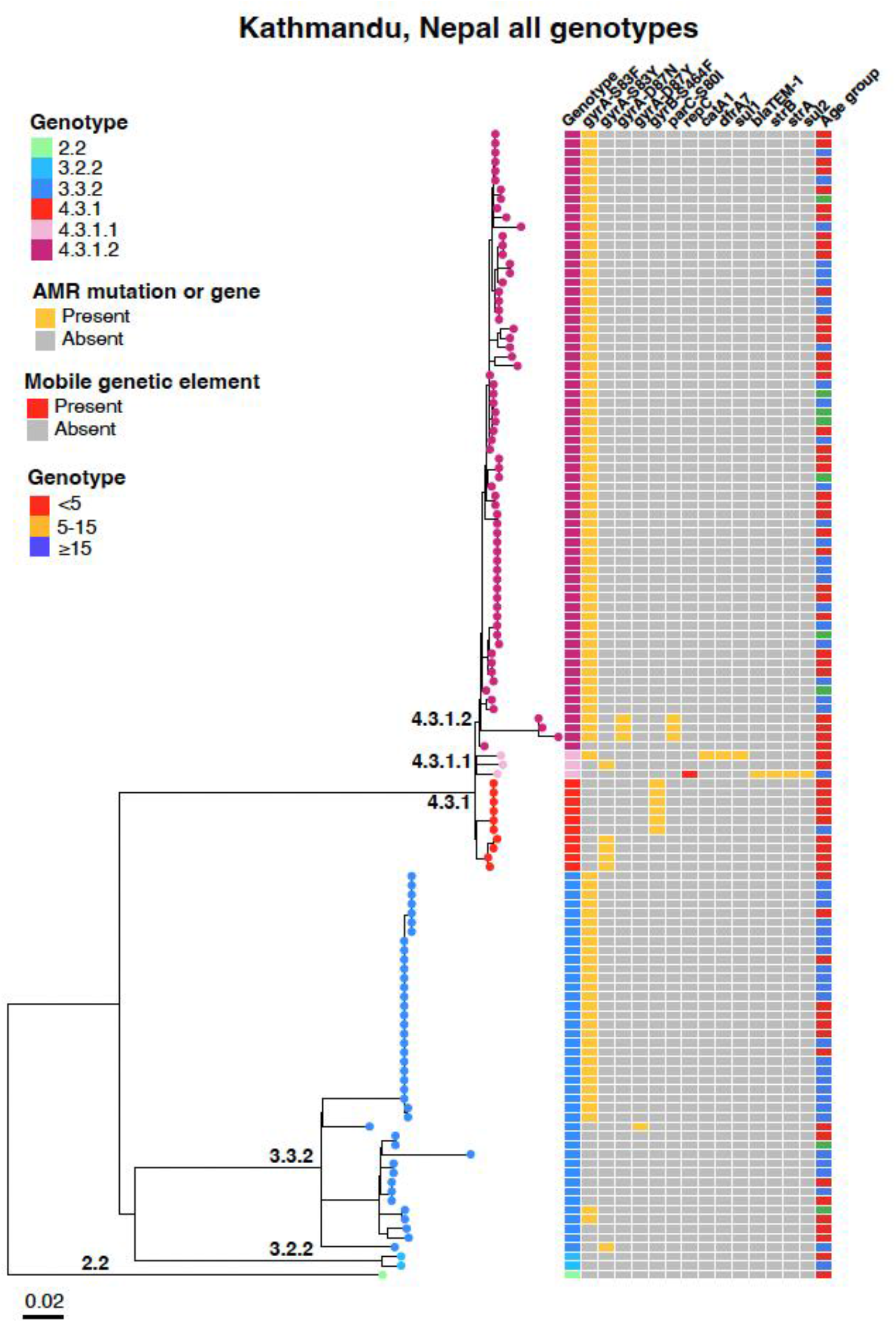
Phylogenetic tree of *S.* Typhi sequenced from STRATAA surveillance in Kathmandu. Whole-genome maximum-likelihood phylogeny of n=124 *S.* Typhi sequences from Kathmandu, Nepal (main ‘STRATAA’ dataset). Tip colours and first column of heatmap indicate the genotype of the sequenced isolate (as per inset legend and branch labels). ‘Columns 2-7 indicate presence of quinolone resistance-associated mutations in *gyrA* and *parC*, remaining columns indicate presence of *repC* (a marker of the common multidrug resistant transposon), acquired genes associated with resistance to first-line drugs, and patient age group (coloured as per inset legends). Branch lengths indicate substitutions per variable site, as per inset scalebar. Interactive phylogeny available at: https://microreact.org/project/nJ42r2XHmd7gZCQ1eqQHGT.

### Supplementary Tables

**Table S1:** *S.* Typhi and *S*. Paratyphi A genomes sequenced in this study

[CSV file]

**Table S2.** Logistic regression models for association of age and sex with pathogen features. Multivariable logistic regression models were fit separately for each country and outcome variable, with age group and sex as predictors. The total number (n) of individuals with each outcome variable, included in each model, is indicated. Age group was treated as an ordinal categorical variable: <5 years, ≥5 and <15 years, ≥15 years. OR, odds ratio; CI, confidence interval; *p-value <0.05. As MDR pathogens were absent in Nepal and almost universal in Malawi, we did not assess association with age in these locations; similarly for ciprofloxacin susceptibility, which was almost invariant within sites, see **Fig. S1c**.

| Country | Modelled Outcome | Predictor | p-value | OR (95% CI) |
| --- | --- | --- | --- | --- |
| Bangladesh | **Serovar**<br>*S. Paratyphi* A (n=95)<br>vs *S. Typhi* (n=357) | Age group | 0.0001* | 1.93 (1.39, 2.71) |
|  |  | Sex (male) | 0.20 | 1.36 (0.86, 2.18) |
|  | **S. Typhi Genotype**<br>Non-H58 *S. Typhi* (n=44)<br>vs H58 *S. Typhi* (n=80) | Age group | 0.01* | 1.45 (1.08, 1.95) |
|  |  | Sex (male) | 0.33 | 1.23 (0.81, 1.88) |
|  | **S. Paratyphi A Genotype**<br>2.4.4 *S. Paratyphi* A (n=56)<br>vs other *S. Paratyphi* A (n=39) | Age group | 0.57 | 0.84 (0.46, 1.52) |
|  |  | Sex (male) | 1.0 | 1.0 (0.43, 2.30) |
|  | **S. Typhi Multidrug Resistance**<br>MDR *S. Typhi* (n=143)<br>vs non-MDR *S. Typhi* (n=214) | Age group | 0.02* | 0.70 (0.52, 0.94) |
|  |  | Sex (male) | 0.70 | 0.92 (0.60, 1.41) |
| Nepal | **Serovar**<br>*S. Paratyphi* A (n=14)<br>vs *S. Typhi* (n=124) | Age group | 0.18 | 0.56 (0.24, 1.34) |
|  |  | Sex (male) | 0.62 | 1.34 (0.43, 4.64) |
|  | **S. Typhi Genotype**<br>Non-H58 *S. Typhi* (n=261)<br>vs H58 *S. Typhi* (n=191) | Age group | 0.15 | 0.65 (0.36, 1.17) |
|  |  | Sex (male) | 0.35 | 1.45 (0.68, 3.17) |
|  | **S. Paratyphi A Genotype**<br>2.4.3 *S. Paratyphi* A (n=8)<br>vs other *S. Paratyphi* A (n=3) | Age group | 0.81 | 0.78 (0.09, 5.89) |
|  |  | Sex (male) | 0.92 | 1.12 (0.11, 12.7) |
| Malawi | **S. Typhi Genotype**<br>Non-H58 *S. Typhi* (n=3)<br>vs H58 *S. Typhi* (n=138) | Age group | 0.79 | 0.80 (0.14, 4.46) |
|  |  | Sex (male) | 1.0 | 0.0 (n/a) |

**Table S3.**
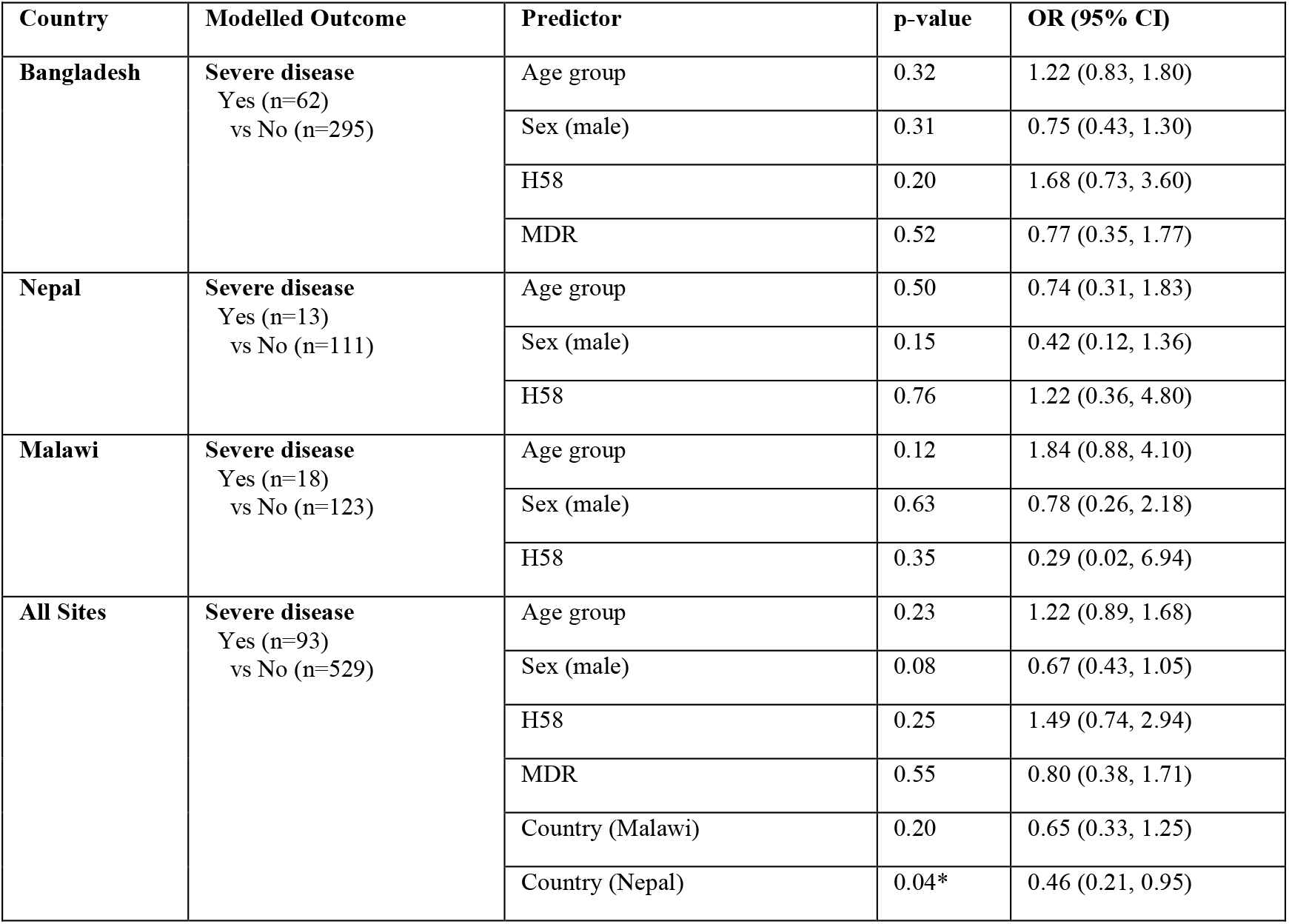
Logistic regression models of severe *S.* Typhi infection. Multivariable logistic regression models were fit separately for each country, and a combined model including all sites as indicated. Age group, sex and H58 genotype were predictors in all models. Multidrug resistance (MDR) was included in the Bangladesh model as it varied across the population and with age, but was excluded from Nepal and Malawi models as it is invariant (near-universal in Malawi, absent in Nepal; see **Fig. S1b**). Quinolone resistance determining region (QRDR) mutation status was excluded from individual site models as it was invariant within sites; and from the all-sites model as it is confounded with country (see **Fig. S1c**). The total number (n) of individuals with each outcome variable, included in each model, is indicated. Age group was treated as an ordinal categorical variable: <5 years, ≥5 and <15 years, ≥15 years. OR, odds ratio; CI, confidence interval; *p-value <0.05.

| Country | Modelled Outcome | Predictor | p-value | OR (95% CI) |
| --- | --- | --- | --- | --- |
| Bangladesh | Severe disease<br>Yes (n=62)<br>vs No (n=295) | Age group | 0.32 | 1.22 (0.83, 1.80) |
|  |  | Sex (male) | 0.31 | 0.75 (0.43, 1.30) |
|  |  | H58 | 0.20 | 1.68 (0.73, 3.60) |
|  |  | MDR | 0.52 | 0.77 (0.35, 1.77) |
| Nepal | Severe disease<br>Yes (n=13)<br>vs No (n=111) | Age group | 0.50 | 0.74 (0.31, 1.83) |
|  |  | Sex (male) | 0.15 | 0.42 (0.12, 1.36) |
|  |  | H58 | 0.76 | 1.22 (0.36, 4.80) |
| Malawi | Severe disease<br>Yes (n=18)<br>vs No (n=123) | Age group | 0.12 | 1.84 (0.88, 4.10) |
|  |  | Sex (male) | 0.63 | 0.78 (0.26, 2.18) |
|  |  | H58 | 0.35 | 0.29 (0.02, 6.94) |
| All Sites | Severe disease<br>Yes (n=93)<br>vs No (n=529) | Age group | 0.23 | 1.22 (0.89, 1.68) |
|  |  | Sex (male) | 0.08 | 0.67 (0.43, 1.05) |
|  |  | H58 | 0.25 | 1.49 (0.74, 2.94) |
|  |  | MDR | 0.55 | 0.80 (0.38, 1.71) |
|  |  | Country (Malawi) | 0.20 | 0.65 (0.33, 1.25) |
|  |  | Country (Nepal) | 0.04* | 0.46 (0.21, 0.95) |

**Table S4.** Comparison of phenotypic and genotypic assessment of non-susceptibility to antimicrobials for *S.* Typhi. Drugs for which phenotype results were available for ≥20% of isolates are shown. Positive predictive value (PPV) and negative predictive value (NPV) of genetic determinants for prediction of resistant phenotypes are shown. Genetic determinants used as predictors of resistance are listed in Supplementary Methods.

| Drug class | No. tested | Resistant phenotype<br>N (% of total tested) | Resistant genotype<br>N (% of total tested) | PPV | NPV |
| --- | --- | --- | --- | --- | --- |
| **Beta-lactamases** |  |  |  |  |  |
| Ampicillin | 575 | 287 (50%) | 286 (50%) | 99.7% | 99.7% |
| Ceftriaxone | 538 | 0 | 0 | - | 100% |
| Cefixime | 357 | 0 | 0 | - | 100% |
| **Folate pathway inhibitors** |  |  |  |  |  |
| Co-trimoxazole | 552 | 304 (55%) | 302 (55%) | 99.7% | 99.2% |
| **Carbapenems** |  |  |  |  |  |
| Meropenem | 354 | 0 | 0 | - | 100% |
| **Macrolides** |  |  |  |  |  |
| Azithromycin | 469 | 13 (2.8%) | 2 (0.4%) | 50% | 97.4% |
| **Chloramphenicol** |  |  |  |  |  |
| Chloramphenicol | 607 | 305 (50%) | 301 (50%) | 98.7% | 98.7% |
| **Quinolones** |  |  |  |  |  |
| Nalidixic Acid | 444 | 437 (98%) | 7 (1.6%) | 100% | 98.4% |
| **Fluoroquinolones** |  |  |  |  |  |
| Ciprofloxacin | 611 | 450 (74%) | 444 (73%) | 96.5% | 96.0% |

**Table S5.** Comparison of phenotypic and genotypic assessment of non-susceptibility to antimicrobials for *S.* Paratyphi. **A.** Drugs for which phenotype results were available for ≥20% of isolates are shown. Positive predictive value (PPV) and negative predictive value (NPV) of genetic determinants for prediction of resistant phenotypes are shown. Genetic determinants used as predictors of resistance are listed in Supplementary Methods.

| Drug class | No. tested | Resistant phenotype<br>N (% of total tested) | Resistant genotype<br>N (% of total tested) | PPV | NPV |
| --- | --- | --- | --- | --- | --- |
| **Beta-lactamases** |  |  |  |  |  |
| Ampicillin | 104 | 0 | 0 | - | 100% |
| Ceftriaxone | 99 | 0 | 0 | - | 100% |
| Cefixime | 94 | 0 | 0 | - | 100% |
| **Folate pathway inhibitors** |  |  |  |  |  |
| Co-trimoxazole | 103 | 0 | 0 | - | 100% |
| **Carbapenems** |  |  |  |  |  |
| Meropenem | 93 | 0 | 0 | - | 100% |
| **Macrolides** |  |  |  |  |  |
| Azithromycin | 109 | 44 (40%) | 3 (2.8%) | 66.7% | 60.4% |
| **Chloramphenicol** |  |  |  |  |  |
| Chloramphenicol | 109 | 0 | 0 | - | 100% |
| **Quinolones** |  |  |  |  |  |
| Nalidixic Acid | 109 | 109 (100%) | 109 (100%) | 100% | - |
| **Fluoroquinolones** |  |  |  |  |  |
| Ciprofloxacin | 109 | 108 (99%) | 109 (100%) | 99.1% | - |

**Table S6:** Publicly available *S.* Typhi and *S*. Paratyphi A genome sequences used to provide global context for genotypes detected in STRATAA sequences

[CSV file]

**Table S7:** Representative *S.* Typhi sequences (one per defined genotype) used to outgroup-root *S*. Typhi phylogenies

[CSV file]

## References

1 GBD 2017 Typhoid and Paratyphoid Collaborators. The global burden of typhoid and paratyphoid fevers: a systematic analysis for the Global Burden of Disease Study 2017. Lancet Infec Dis 2019; 19: 369–81.

2 Rahman SIA, Dyson ZA, Klemm EJ, et al. Population structure and antimicrobial resistance patterns of Salmonella Typhi isolates in urban Dhaka, Bangladesh from 2004 to 2016. PLoS Negl Trop Dis 2020; 14: e0008036.

3 Britto CD, Dyson ZA, Duchene S, et al. Laboratory and molecular surveillance of paediatric typhoidal Salmonella in Nepal: Antimicrobial resistance and implications for vaccine policy. PLoS Negl Trop Dis 2018; 12: e0006408.

4 Klemm EJ, Shakoor S, Page AJ, et al. Emergence of an Extensively Drug-Resistant Salmonella enterica Serovar Typhi Clone Harboring a Promiscuous Plasmid Encoding Resistance to Fluoroquinolones and Third-Generation Cephalosporins. mBio 2018; 9. DOI:10.1128/mbio.00105-18.

5 Hooda Y, Sajib MSI, Rahman H, et al. Molecular mechanism of azithromycin resistance among typhoidal Salmonella strains in Bangladesh identified through passive pediatric surveillance. PLoS Negl Trop Dis 2019; 13: e0007868.

6 Duy PT, Dongol S, Giri A, et al. The emergence of azithromycin-resistant Salmonella Typhi in Nepal. JAC Antimicrob Resist 2020; 2: dlaa109.

7 Silva KE da, Tanmoy AM, Pragasam AK, et al. The international and intercontinental spread and expansion of antimicrobial-resistant Salmonella Typhi: a genomic epidemiology study. Lancet Microbe 2022; 3: e567–77.

8 Wong VK, Baker S, Pickard DJ, et al. Phylogeographical analysis of the dominant multidrug-resistant H58 clade of Salmonella Typhi identifies inter- and intracontinental transmission events. Nat Genet 2015; 47: 632–9.

9 Feasey NA, Gaskell K, Wong V, et al. Rapid emergence of multidrug resistant, H58- lineage Salmonella typhi in Blantyre, Malawi. PLoS Negl Trop Dis 2015; 9: e0003748.

10 Thanh DP, Karkey A, Dongol S, et al. A novel ciprofloxacin-resistant subclade of H58 Salmonella Typhi is associated with fluoroquinolone treatment failure. eLife 2016; 5: e14003.

11 Birkhold M, Mwisongo A, Pollard AJ, Neuzil KM. Typhoid Conjugate *Vaccine*s: Advancing the Research and Public Health Agendas. J Infect Dis 2021; 224: S781–7.

12 Nampota-Nkomba N, Nyirenda OM, Khonde L, et al. Safety and immunogenicity of a typhoid conjugate vaccine among children aged 9 months to 12 years in Malawi: a nested substudy of a double-blind, randomised controlled trial. Lancet Glob Health 2022; 10: e1326–35.

13 Shakya M, Colin-Jones R, Theiss-Nyland K, et al. Phase 3 Efficacy Analysis of a Typhoid Conjugate Vaccine Trial in Nepal. N Engl J Med 2019; 381: 2209–18.

14 Patel PD, Patel P, Liang Y, et al. Safety and Efficacy of a Typhoid Conjugate Vaccine in Malawian Children. NJEM 2021; 385: 1104–15.

15 Qadri F, Khanam F, Liu X, et al. Protection by vaccination of children against typhoid fever with a Vi-tetanus toxoid conjugate vaccine in urban Bangladesh: a cluster-randomised trial. Lancet 2021; 398: 675–84.

16 Yousafzai MT, Karim S, Qureshi S, et al. Effectiveness of typhoid conjugate vaccine against culture-confirmed Salmonella enterica serotype Typhi in an extensively drug-resistant outbreak setting of Hyderabad, Pakistan: a cohort study. Lancet Glob Health 2021; 9: e1154–62.

17 Poncin M, Marembo J, Chitando P, et al. Implementation of an outbreak response vaccination campaign with typhoid conjugate vaccine – Harare, Zimbabwe, 2019. Vaccine X 2022; 12: 100201.

18 Thobani RS, Yousafzai MT, Sultana S, et al. Field evaluation of typhoid conjugate vaccine in a catch-up campaign among children aged 9 months to 15 years in Sindh, Pakistan. Vaccine 2022; 40: 5391–8.

19 Shakya M, Neuzil KM, Pollard AJ. Prospects of future typhoid and paratyphoid vaccines in endemic countries. J Infect Dis 2021; 224: jiab393.

20 Meiring JE, Shakya M, Khanam F, et al. Burden of enteric fever at three urban sites in Africa and Asia: a multicentre population-based study with 626,219 person-years of observation. Lancet Infect Dis 2021.

21 Rahman SIA, Nguyen TNT, Khanam F, et al. Genetic diversity of Salmonella Paratyphi A isolated from enteric fever patients in Bangladesh from 2008 to 2018. Plos Neglect Trop D 2021; 15: e0009748.

22 Darton TC, Meiring JE, Tonks S, et al. The STRATAA study protocol: a programme to assess the burden of enteric fever in Bangladesh, Malawi and Nepal using prospective population census, passive surveillance, serological studies and healthcare utilisation surveys. BMJ Open 2017; 7: e016283.

23 Skittrall JP, Levy D, Obichukwu C, et al. Azithromycin susceptibility testing of Salmonella enterica serovar Typhi: impact on management of enteric fever. Clin Infect Pract 2021; 10: 100069.

24 Campbell F, Strang C, Ferguson N, Cori A, Jombart T. When are pathogen genome sequences informative of transmission events? PLoS Pathog 2018; 14: e1006885–21.

25 Tanmoy AM, Westeel E, Bruyne KD, et al. Salmonella enterica Serovar Typhi in Bangladesh: Exploration of Genomic Diversity and Antimicrobial Resistance. mBio 2018; 9: e02112–18.

26 Gauld JS, Olgemoeller F, Heinz E, et al. Spatial and genomic data to characterize endemic typhoid transmission. Clin Infect Dis 2022; 74: ciab745-.

27 Kariuki S, Dyson ZA, Mbae C, et al. Multiple introductions of multidrug-resistant typhoid associated with acute infection and asymptomatic carriage, Kenya. eLife 2021; 10: e67852.

28 Manson AL, Cohen KA, Abeel T, et al. Genomic analysis of globally diverse Mycobacterium tuberculosis strains provides insights into the emergence and spread of multidrug resistance. Nat Genet 2017; 49: 395–402.

29 Duchene S, Holt KE, Weill F-X, et al. Genome-scale rates of evolutionary change in bacteria. Microb Genom 2016; 2: e000094.

30 Dyson ZA, Holt KE. Five years of GenoTyphi: updates to the global Salmonella Typhi genotyping framework. J Infect Dis 2021; 224: jiab414.

## Supplementary References

1 Wong VK, Baker S, Pickard DJ, et al. Phylogeographical analysis of the dominant multidrug-resistant H58 clade of Salmonella Typhi identifies inter- and intracontinental transmission events. Nat Genet 2015; 47: 632–9.

2 Parkhill J, Dougan G, James KD, et al. Complete genome sequence of a multiple drug resistant Salmonella enterica serovar Typhi CT18. Nature 2001; 413: 848–52.

3 Holt KE, Thomson NR, Wain J, et al. Pseudogene accumulation in the evolutionary histories of Salmonella enterica serovars Paratyphi A and Typhi. BMC Genomics 2009; 10: 36.

4 Langmead B, Salzberg SL. Fast gapped-read alignment with Bowtie 2. Nat Meth 2012; 9: 357–9.

5 Li H, Durbin R. Fast and accurate short read alignment with Burrows–Wheeler transform. Bioinformatics 2009; 25: 1754–60.

6 Britto CD, Dyson ZA, Duchene S, et al. Laboratory and molecular surveillance of paediatric typhoidal Salmonella in Nepal: Antimicrobial resistance and implications for vaccine policy. PLoS Negl Trop Dis 2018; 12: e0006408.

7 Dyson ZA, Holt KE. Five Years of GenoTyphi: Updates to the Global Salmonella Typhi Genotyping Framework. J Infect Dis 2021; 224: S775–80.

8 Wong VK, Baker S, Connor TR, et al. An extended genotyping framework for Salmonella enterica serovar Typhi, the cause of human typhoid. Nat Commun 2016; 7: 12827.

9 Tanmoy AM, Hooda Y, Sajib MSI, et al. Paratype: A genotyping framework and an open-source tool for Salmonella Paratyphi A. Medrxiv 2021; : 2021.11.13.21266165.

10 Britto CD, Dyson ZA, Mathias S, et al. Persistent circulation of a fluoroquinolone-resistant Salmonella enterica Typhi clone in the Indian subcontinent. J Antimicrob Chemother 2020; 75: 337–41.

11 Tanmoy AM, Westeel E, Bruyne KD, et al. Salmonella enterica Serovar Typhi in Bangladesh: Exploration of Genomic Diversity and Antimicrobial Resistance. mBio 2018; 9: e02112–18.

12 Dyson ZA, Thanh DP, Bodhidatta L, et al. Whole Genome Sequence Analysis of Salmonella Typhi Isolated in Thailand before and after the Introduction of a National Immunization Program. PLoS Negl Trop Dis 2017; 11: e0005274.

13 Klemm EJ, Shakoor S, Page AJ, et al. Emergence of an Extensively Drug-Resistant Salmonella enterica Serovar Typhi Clone Harboring a Promiscuous Plasmid Encoding Resistance to Fluoroquinolones and Third-Generation Cephalosporins. mBio 2018; 9. DOI:10.1128/mbio.00105-18.

14 Matono T, Morita M, Yahara K, et al. Emergence of Resistance Mutations in Salmonella enterica Serovar Typhi Against Fluoroquinolones. Open Forum Infect Dis 2017; 4: ofx230.

15 Pragasam AK, Pickard D, Wong V, et al. Phylogenetic Analysis Indicates a Longer Term Presence of the Globally Distributed H58 Haplotype of Salmonella Typhi in Southern India. Clin Infect Dis 2020; 71: 1856–63.

16 Rahman SIA, Dyson ZA, Klemm EJ, et al. Population structure and antimicrobial resistance patterns of Salmonella Typhi isolates in urban Dhaka, Bangladesh from 2004 to 2016. PLoS Negl Trop Dis 2020; 14: e0008036.

17 Rasheed F, Saeed M, Alikhan N-F, et al. Emergence of Resistance to Fluoroquinolones and Third-Generation Cephalosporins in Salmonella Typhi in Lahore, Pakistan. Microorganisms 2020; 8: 1336.

18 Thanh DP, Karkey A, Dongol S, et al. A novel ciprofloxacin-resistant subclade of H58 Salmonella Typhi is associated with fluoroquinolone treatment failure. eLife 2016; 5: e14003.

19 International Typhoid Consortium, Wong VK, Holt KE, et al. Molecular Surveillance Identifies Multiple Transmissions of Typhoid in West Africa. PLoS Negl Trop Dis 2016; 10: e0004781.

20 Holt KE, Parkhill J, Mazzoni CJ, et al. High-throughput sequencing provides insights into genome variation and evolution in Salmonella Typhi. Nat Genet 2008; 40: 987–93.

21 Ingle DJ, Nair S, Hartman H, et al. Informal genomic surveillance of regional distribution of Salmonella Typhi genotypes and antimicrobial resistance via returning travellers. PLoS Negl Trop Dis 2019; 13: e0007620.

22 Croucher NJ, Page AJ, Connor TR, et al. Rapid phylogenetic analysis of large samples of recombinant bacterial whole genome sequences using Gubbins. Nucleic Acids Research 2015; 43: e15–e15.

23 Stamatakis A. RAxML version 8: a tool for phylogenetic analysis and post-analysis of large phylogenies. Bioinformatics 2014; 30: 1312–3.

24 Argimon S, Abudahab K, Goater RJE, et al. Microreact: visualizing and sharing data for genomic epidemiology and phylogeography. Microbial Genomics 2016; 2: e000093.

25 Yu G, Smith DK, Zhu H, Guan Y, Lam TT-Y. ggtree: an rpackage for visualization and annotation of phylogenetic trees with their covariates and other associated data. Methods Ecol Evol 2016; 8: 28–36.

26 Hooda Y, Sajib MSI, Rahman H, et al. Molecular mechanism of azithromycin resistance among typhoidal Salmonella strains in Bangladesh identified through passive pediatric surveillance. PLoS Negl Trop Dis 2019; 13: e0007868.

27 Argimon S, Yeats CA, Goater RJ, et al. A global resource for genomic predictions of antimicrobial resistance and surveillance of Salmonella Typhi at pathogenwatch. Nat Commun 2021; 12: 2879–12.

28 Inouye M, Dashnow H, Raven L-A, et al. SRST2: Rapid genomic surveillance for public health and hospital microbiology labs. Genome Med 2014; 6: 90.

29 Gupta SK, Padmanabhan BR, Diene SM, et al. ARG-ANNOT, a new bioinformatic tool to discover antibiotic resistance genes in bacterial genomes. Antimicrob Agents Chemother 2014; 58: 212–20.

30 Carattoli A, Zankari E, García-Fernández A, et al. In silico detection and typing of plasmids using PlasmidFinder and plasmid multilocus sequence typing. Antimicrob Agents Chemother 2014; 58: 3895–903.

31 Chattaway MA, Gentle A, Nair S, et al. Phylogenomics and antimicrobial resistance of Salmonella Typhi and Paratyphi A, B and C in England, 2016–2019. Microb Genom 2021; 7: 000633.

32 Wilm A, Aw PPK, Bertrand D, et al. LoFreq: a sequence-quality aware, ultra-sensitive variant caller for uncovering cell-population heterogeneity from high-throughput sequencing datasets. Nucleic Acids Research 2012; 40: 11189–201.

33 Knaus BJ, Grünwald NJ. vcfr: a package to manipulate and visualize variant call format data in R. Molecular Ecology Resources 2017; 17: 44–53.

34 Jaillard M, Lima L, Tournoud M, et al. A fast and agnostic method for bacterial genome-wide association studies: Bridging the gap between k-mers and genetic events. PLoS Genet 2018; 14: e1007758.

35 Wick RR, Judd LM, Gorrie CL, Holt KE. Unicycler: Resolving bacterial genome assemblies from short and long sequencing reads. PLoS Comput Biol 2017; 13: e1005595.

36 Schliep KP. phangorn: phylogenetic analysis in R. Bioinformatics 2011; 27: 592–3.

37 Dixon P. VEGAN, a package of R functions for community ecology. Journal of Vegetaton Sciences 2003; 14: 927–30.

38 Kembel SW, Cowan PD, Helmus MR, et al. Picante: R tools for integrating phylogenies and ecology. Bioinformatics 2010; 26: 1463–4.

39 Blomberg SP, Garland T, Ives AR. Testing for phylogenetic signal in comparative data: behavioral traits are more labile. Evolution 2003; 57: 717–45.

40 Silva KE da, Tanmoy AM, Pragasam AK, et al. The international and intercontinental spread and expansion of antimicrobial-resistant Salmonella Typhi: a genomic epidemiology study. Lancet Microbe 2022; 3: e567–77.

